# Characterization of a fieldable process for airborne virus detection

**DOI:** 10.1101/2023.07.03.23292170

**Authors:** Huifeng Du, Simone Bruno, Kalon J. Overholt, Sebastian Palacios, Hsin-Ho Huang, Carlos Barajas, Ben Gross, Cindy Lee, Haley K. Evile, Nuno Rufino de Sousa, Antonio Gigliotti Rothfuchs, Domitilla Del Vecchio

**Author notes:** These authors contributed equally: H. Du, S. Bruno, K.J. Overholt, and S. Palacios.

## Abstract

Rapid, on-site, airborne virus detection is a requirement for timely action against the spread of air-transmissible infectious diseases. This applies both to future threats and to common viral diseases, such as influenza and COVID-19, which hit vulnerable populations yearly with severe consequences. The ultra-low concentrations of virus in the air make airborne virus detection difficult, yet readily infect individuals when breathed. Here, we propose a fieldable process that includes an enrichment step to concentrate collected genetic material in a small volume. The enrichment approach uses capillary electrophoresis and an RT-qPCR-compatible buffer, which allow enrichment of the RNA by about 5-fold within only 10 minutes of operation. Our detection process consists of air sampling through electrostatic precipitation, RNA extraction via heating, RNA enrichment, and RT-qPCR for detection. We optimized each step of the process and estimated a detection sensitivity of 3106 *±* 2457 genome copies (gc) per m^3^ of air. We then performed an integration experiment and confirmed a sensitivity of 5654 gc/m^3^ with a detection rate of 100% and a sensitivity of 4221 gc/m^3^ with a detection rate of 78.6%. When using fast RT-qPCR, the latency of the whole process is down to 61 minutes. Given that our sensitivity falls in the low range of influenza and SARS-CoV-2 concentrations reported in indoor spaces, our study shows that, with enrichment, airborne pathogen detection can be made sufficiently sensitive for practical use.

## 1 Introduction

The COVID-19 pandemic has demonstrated how vulnerable we are to the spread of airborne infectious diseases. It is also not an isolated case, but rather it follows a series of other epidemics, although not as devastating, including SARS (2003), Swine Flu (2009), MERS (2012), Ebola (2014), and the 2018 United States adenovirus outbreak [1, 2, 3, 4, 5]. With a major epidemic every 2-6 years, we are at continuous risk for the spread of novel infectious diseases. At the same time, even common viruses, such as influenza, RSV, and now also SARS-CoV-2, have major repercussions every year among vulnerable populations, leaving behind deaths and healthcare expenses [6, 7, 8].

Early detection is required for timely action and acting quickly has been proven one of the most effective means to contain an epidemic [9, 10, 11]. Currently, despite the numerous COVID-19 surveillance programs, there is an average delay of 11 days between the onset of illness in patients and their diagnosis and reporting on infection curves [12]. Standard detection approaches typically rely on symptom-based viral testing, at which point an ill individual has already had the time to spread the virus [13]. Even routine PCR testing has turnaround times of about 24 hours or more [14, 15], and is also not financially feasible for many institutions. Environmental monitoring approaches that sample the air of indoor spaces and rapidly analyze it for the presence of pathogens is a more practical approach that could reduce costs, improve detection speed, and minimize disruption to schedules. Since timely actions can only be taken if detection is sufficiently fast and sensitive, the speed and sensitivity of detection are two critical features of an on-site airborne virus detection system.

Currently, there are no established processes for early detection of airborne pathogens, such as viruses and bacteria, in the air of indoor environments. Existing technologies either fail to provide timely information or lack the necessary sensitivity to minimize false negative readings. Common approaches include sampling the air with a pump and then processing the sample in a laboratory, which takes days (BioWatch program [16] and similar [17]). On-site bio-sensing technologies have appeared that could reportedly detect the presence of SARS-CoV-2, but only at concentrations as high as 31.8 *·* 10^3^ genome copy/m^3^ (gc*/*m^3^) or 31.8 gc*/*L [18]. This value is among the highest concentrations of influenza virus during flu season and SARS-CoV-2 in COVID-19 hospital patient rooms, reported to be as low as 5 *·* 10^3^ gc*/*m^3^ or 5 gc*/*L [17, 19]. Although low, these concentrations can cause illness with high probability if breathed for more than 1 hour [20]. Therefore, these methods are not sufficiently sensitive to inform actions that could make a difference. Insufficient sensitivity also plagues the Thermo Scientific’s AeroSense Sampler and Bertin Technologies’ Coriolis µ, with their separate PCR processes, which can detect common respiratory pathogens, including SARS-CoV-2, Flu A/B and/or RSV, and microbes on-site [18, 21, 22]. In summary, the ultra-low concentrations of virus in the air (as low as 1.84 gc*/*L in some cases [23, 24]) when compared to those in nasal swabs of sick patients (10^4^ *−* 10^7^gc/mL [25]) pose a unique challenge to the airborne virus detection problem.

To tackle this challenge, we propose a fieldable airborne virus detection process that incorporates an RNA enrichment step to concentrate the viral genetic material in a small volume (Fig. 1). The process uses electrostatic precipitation (ESP) for sampling the air, and collects aerosols onto a metal probe. Subsequently, the metal probe is heated in a small volume of water to extract the RNA. Once the RNA is extracted, the enrichment step performs capillary electrophoresis to concentrate the RNA in a small volume, which is then directly used in an RT-qPCR reaction for detection (Fig. 2). Although we use SARS-CoV2 as a test bed, the process that we propose is applicable to any RNA or DNA virus whose genetic sequence is known.

**Figure 1:**
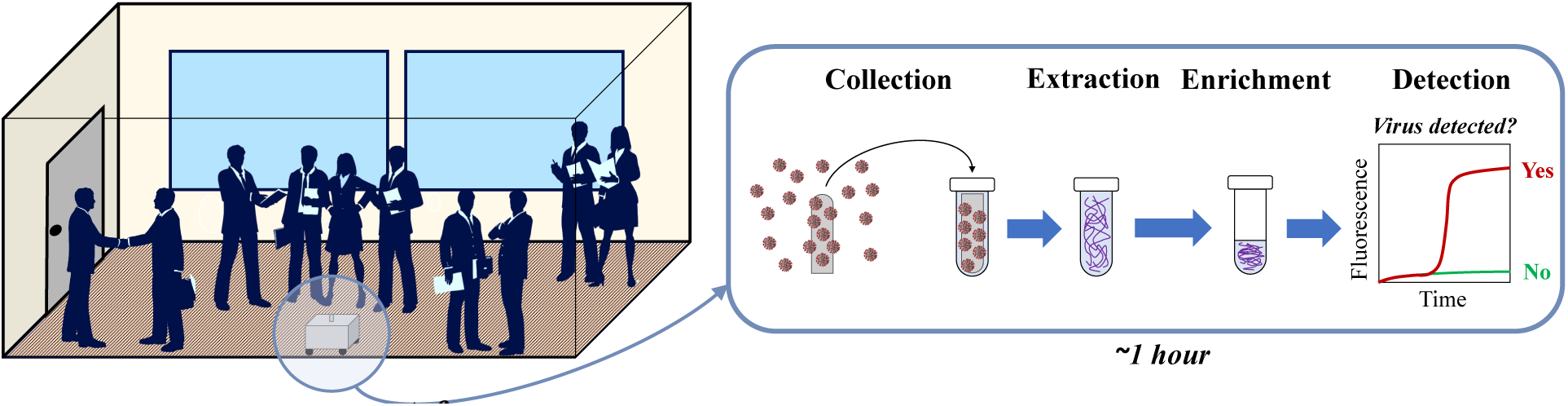
Proposed concept for on-site detection of airborne pathogens. A small platform collects the air through electrostatic precipitation (ESP), extracts and concentrates the genetic material, and then performs chemical detection through RT-qPCR. It outputs information within one hour of operation.

**Figure 2:**
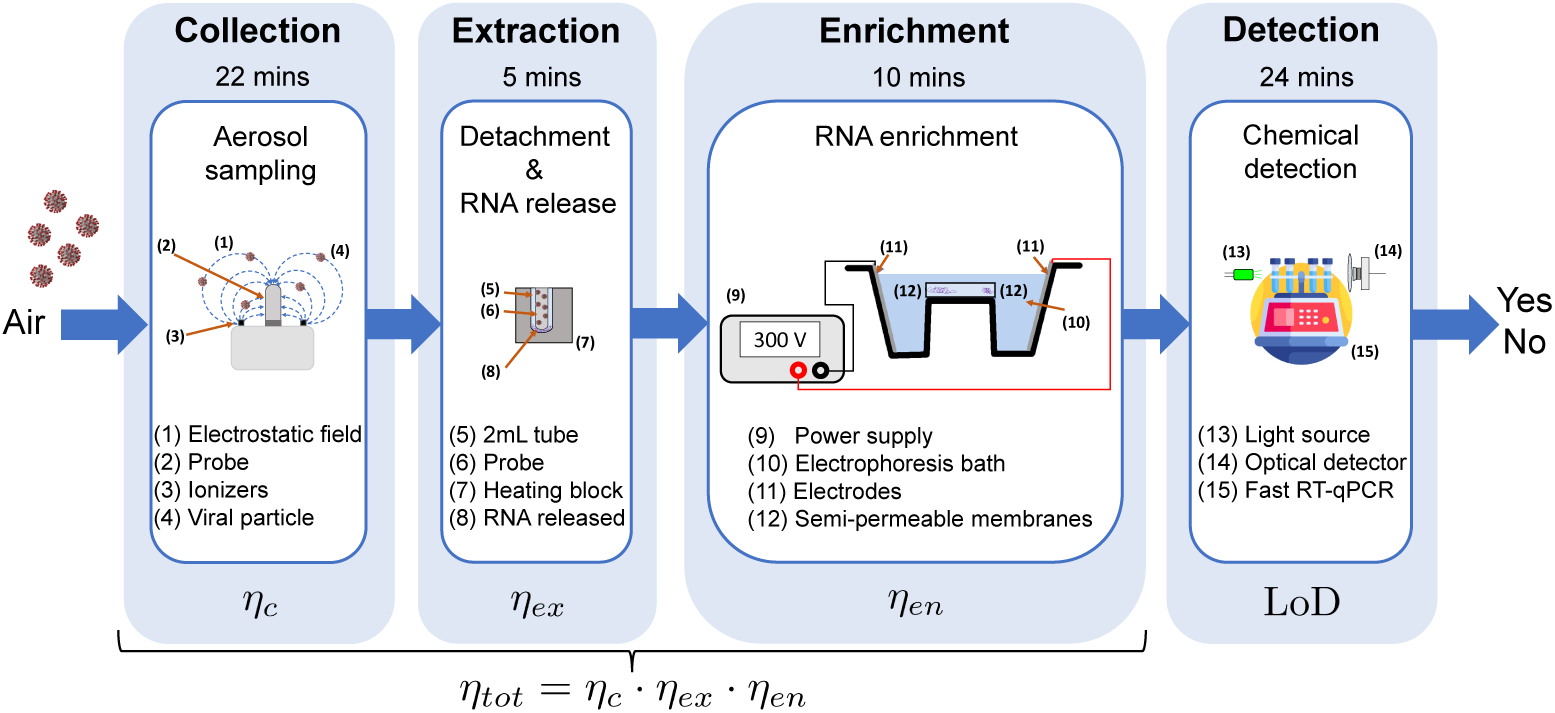
Components of the airborne virus detection process. Air particles are collected on the metal probe of an aerosol sampler, which uses electrostatic precipitation (ESP) and follows the design of [26]. After sampling for 22 minutes, the metal probe with collected particles is placed in a 2 mL tube with 220 µL of nuclease-free water and goes through heating at 98 °C for 5 minutes. The water with the sample is then entirely transferred to an enrichment device, which performs capillary electrophoresis against a semipermeable membrane for 10 minutes. Upon completion, the 7.4 µL of sample next to the membrane is drawn and taken to chemical detection. This employs RT-qPCR with primers specific to the viral target and takes about 24 minutes when using a fast RT-qPCR machine. Each step performance is characterized in terms of its efficiency *η*. The variables *η_*c*_, η_*ex*_,* and *η_*en*_* are the efficiencies of the collection, extraction, and enrichment steps, respectively, as defined in the text, LoD is the limit of detection of RT-qPCR, and *η_*tot*_* represents the efficiency of the overall process.

The paper is organized as follows. In Section 2, we evaluate the performance of all the process’ steps: Collection (Section 2.1), Extraction (Section 2.2), Enrichment (Section 2.3), and Detection (Section 2.4). In Section 3, we perform integration experiments to determine the sensitivity of the entire detection process. Conclusions and discussion are presented in Section 4.

## 2 Performance evaluation of the airborne virus detection process

The airborne virus detection process is depicted in Fig.2. Collection occurs through an air sampler that uses electrostatic precipitation (ESP) and follows the design of [26]. The electrostatic field collects ionized air particles on a metal probe, which is taken to the extraction step after 22 minutes of sampling. Extraction of genetic material is performed by heating the probe submersed in 220 µL water in a 2mL tube at 98 °C for 5 minutes. The entire volume is then injected into a 250 µL channel whose ends are closed by a semipermeable membrane, and then taken to the electrophoresis bath. After 10 minutes of capillary electrophoresis, a 7.4 µL volume is extracted next to the positive electrode, where all the RNA has migrated. This volume is then taken to a 20 µL RT-qPCR reaction for detection. When using a fast RT-qPCR machine [27], chemical detection can be performed in 24 minutes or less.

We evaluate the performance of the whole process by its sensitivity *C_min_*, that is, the lowest concentration of viral particles in the air (in genome copies per m^3^ of air) that can be detected with a detection rate of at least 95%. To this end, we define the efficiency of the whole process *η_tot_* as the ratio between the number of genome copies resulting in the 20 µL RT-qPCR reaction volume and the number of corresponding viral genome copies per m^3^ of air. With *η_tot_*defined this way, and with LoD the limit of detection of RT-qPCR (expressed in cp/µL), the sensitivity is given by

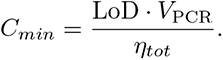

where *V*_PCR_ = 20 µL is the final RT-qPCR reaction volume. We first estimate *C_min_* by experimentally characterizing the efficiency of each of the process’ steps in isolation, and by then calculating *η_tot_* as the product of the efficiencies of each step (Fig. 2). Specifically, we define the collection efficiency *η_c_* as the ratio between the number of particles collected on the metal probe and the number of particles per m^3^ of air. The extraction efficiency *η_ex_* is defined as the ratio between the number of RNA particles extracted in 220 µL of water and the number of viral particles on the metal probe. The enrichment efficiency *η_en_* is the ratio between the number of RNA molecules in the enriched fraction of the volume extracted (7.4 µL) and the number of RNA molecules in the enrichment device volume (250 µL). Then, the overall process efficiency, *η_tot_*, is given by

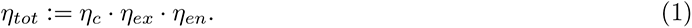

This evaluation of *η_tot_* does not account for potential losses associated with the transfer of the samples through the steps of the process. In order to validate the sensitivity calculated this way while accounting for these losses, we also perform an integration test. In this test, we directly load inactivated virus on the metal probe at copy numbers corresponding to the estimated *C_min_* and evaluate the detection rate.

### 2.1 Characterization of the collection process based on ESP

We characterized the efficiency of aerosol sampling as shown in Fig. 3. Specifically, the objective was to obtain a mapping between the particle concentration in air and the number of particles collected on the ESP metal probe. We opted for a sampler that exploits corona discharge ionization and electrostatic precipitation (ESP). When compared to a pump, the ESP is substantially cheaper, does not produce noise, and only minimally disrupts the airflow, which is an advantage when combining sensing with computational fluid dynamic (CFD) models to estimate particle concentration distributions within a room [28, 29, 30]. Specifically, we selected the portable ESP air sampler developed by De Sousa et al. [26], which is made of a sealed-plastic enclosure (13x13x7.5 cm), with four carbon brush ionizers at the four corners of the upper surface and a magnetic holder in the center of the upper surface. The collector, a stainless-steel metal probe (3.5 cm in length and 0.88 cm in diameter), is magnetically attached to the holder. The ionizers are electrically connected to the negative output voltage of a -20 kV high-voltage supply located within the device, while the magnetic holder is electrically grounded. While in use, the ESP creates a strong electric field between the ionizers and the metal probe, which results in the generation of a corona discharge at the tips of the ionizers. Consequently, electrons are emitted in close proximity to the ionizers’ tips, causing the ionization of airborne particles nearby. The electric field between the ionizers and the collector thus accelerates the ionized particles towards the metal probe, where they collide, lose their charge, and are ultimately collected.

**Figure 3:**
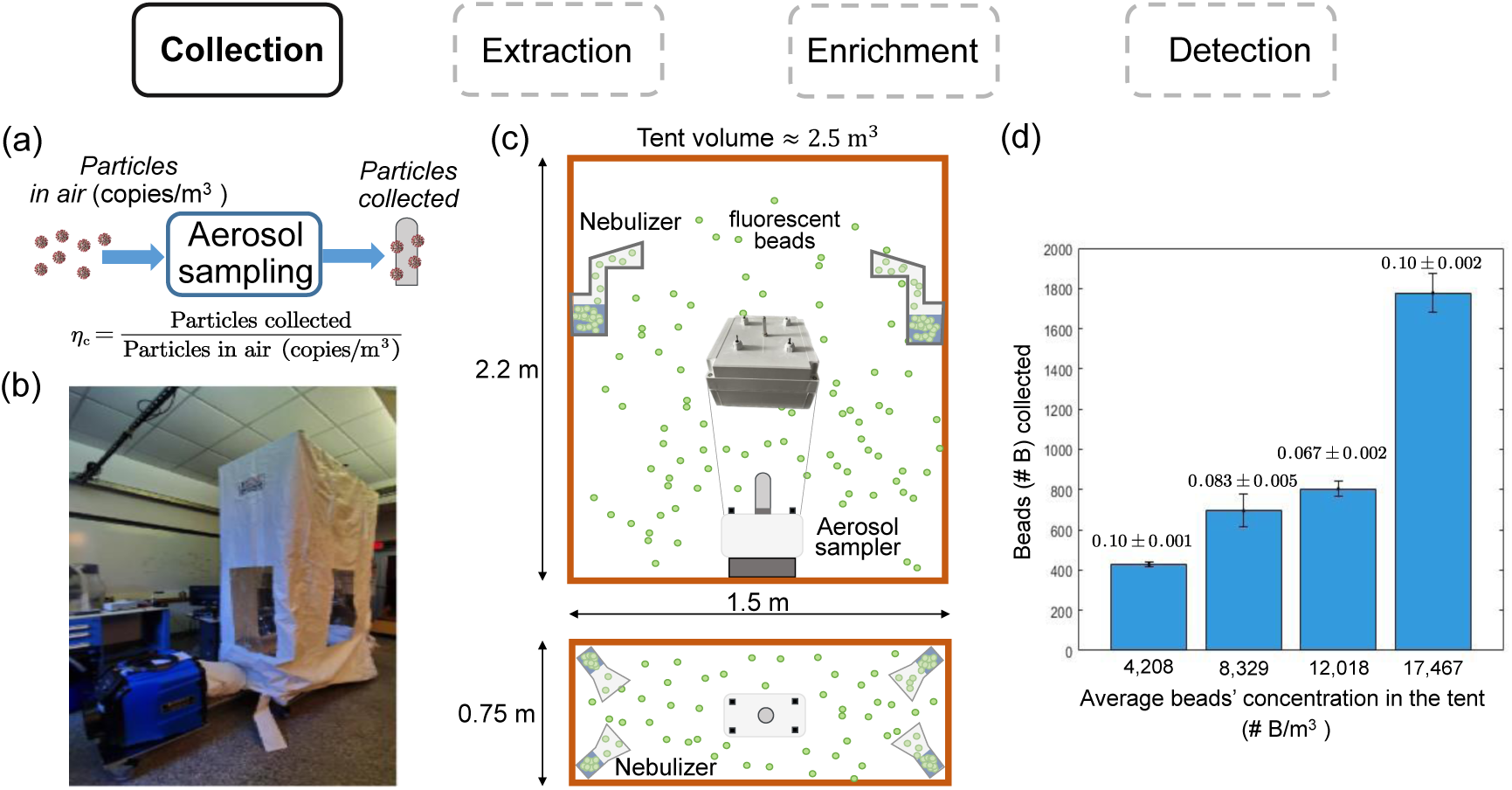
Collection process through ESP. (a) Block diagram. (b) Containment tent where beads were nebulized. (c) Experimental setup: the containment tent is 2.54 m^3^ with the sampler at the center. At the top four corners we locate four nebulizers from which we injected fluorescent beads (B) in the tent. (d) Bar chart showing the number of beads (#*B*) recovered from the metal probe of the ESP device at the end of *T* = 22 min of sampling. On the *x*-axis we indicated the average bead concentrations in the tent 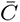 (#*B/m*^3^) and on the *y*-axis the number beads collected. The average and standard deviation is computed based on three experimental replicates using the definition in panel (a). The data was background-subtracted, that is, subtracting the amount of beads collected without nebulization of new particles (see “Methods and Materials” for details).

The experiments were conducted inside a containment tent (Abetement Technologies, AG3000MCCK) (Fig. 3(b),(c)), in which solutions of 1 µm diameter fluorescent beads at varying concentrations (FluoSpheres -Thermo Fisher, F13081) were nebulized. According to Hawks et al. [31], droplets can vary greatly in size, but the smaller ones evaporate into solid particles, and the majority of virus is found within particles smaller than 5 µm of size. Further aerodynamic analysis has reported that the diameter of SARS-CoV-2 aerosols were around 1 µm [32]. We therefore opted for using 1 µm size fluorescent beads, with the understanding that by counting one fluorescent bead as one genome copy of virus may be under-estimating our collection efficiency since each particle likely contains more than one viral copy. In addition, the measurement for the particle size of the Omron nebulizer (NEC801) [33] suggests that most of the nebulized droplets fall within the range of 1-10 µm, with a MMAD (Mass Median Aerodynamic Diameter) of 3 µm. As they evaporate and shrink in size, the size spectrum will continue to shift towards the 1 µm end, consistent with clinical observation of SARS-CoV-2 aerosols [32]. The fluorescent beads were diluted in filtered water to the desired concentrations and introduced in the tent through four Omron nebulizers located at the four top corners of the tent. The aerosol sampler was located in the tent in a central position, 20 cm from the ground and 1.5 m from the nebulizers (Fig. 3(c)). In each experiment, we considered a continuous but small rate of 0.3 mL */* min of nebulization and a simultaneous collection. All experiments lasted *T* = 22 min. Given that our objective was to make the whole detection process run within about one hour, we decreased the sampling time to fit the whole process within the hour time frame, given the duration of all the downstream processes after optimization (Fig. 2).

At the end of sampling, the collector was transferred in a 2 mL microcentrifuge tube and 250 µL of filtered water was added. After vortexing the tube for 1 min, the metal probe was removed thorough a magnet, put in another 2 mL microcentrifuge tube, new filtered water (250 µL) was added, and the tube was again vortexed for 1 min. Then, in order to accurately determine the absolute counts of particles in the samples, a known quantity of counting beads (CountBright Absolute Counting Beads - Invitrogen, C36950) was added to the sample left in each of the two tubes (more details in Methods Section “Use of counting beads for flow cytometry measurements”). By using a BD Accuri C6 flow cytometer (BD Biosciences, San Jose, CA), we quantified the amount of fluorescent beads in all the tubes, representing the total number of fluorescent beads collected on the probe. Before starting each collection experiment, we wiped the surface of the metal probe with the PREempt™RTU disinfectant solution that contains 0.5 % hydrogen peroxide and left it to dry in air for 5 minutes (more details in the Methods section “Metal collector cleaning (0.5% hydrogen peroxide)”). This is because the hydrogen peroxide in the PREempt™solution promotes the generation of free radicals and improves the efficiency of electrostatic precipitators [34, 35].

The results of these experiments are shown in Fig. 3(d). On the *x*-axis we reported the *average bead concentration in the tent* over the sampling time interval *T*, that is 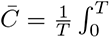 *C*(*t*)*dt*, in which *C*(*t*) represents the instantaneous concentration in the tent at time *t* and *T* = 22 min. The average bead concentrations that we used are consistent with the reported concentrations of influenza virus present during flu season in enclosed environments such as daycare centers and airplanes, ranging from 5,000 to 35,000 copies per *m*^3^ [36]. Specifically, we chose four values of 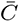 that fall within the lower half of this range. We conclude that the higher 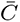, the higher the number of beads collected on the metal probe. Furthermore, the efficiency of collection is *η_c_* = 0.10*±*0.001 at the lowest concentration of 4200 B*/*m^3^ and *η_c_* = 0.083*±*0.005, 0.067*±*0.002, 0.10*±*0.002 for concentrations 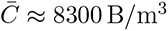, 12 000 B*/*m^3^, 17 500 B*/*m^3^, respectively (Fig. 3(d)).

### 2.2 Characterization of the heat extraction process

Heat extraction has been effectively used in other fieldable biosensing processes as a means to release the genetic material from a virus without the need for additional chemicals [37]. We therefore opted here for a similar approach. Specifically, the metal probe loaded with viral particles is transferred in a 2 mL microcentrifuge tube with 220 µL of nuclease-free water, which is then heated at 98 °C for 5 minutes. For characterizing the efficiency of the heat extraction process, we artificially loaded a known amount of virus on the probe, performed heat extraction, and then transferred 7.4 µL to a 20 µL RT-qPCR reaction.

Specifically, we chose the heat-inactivated SARS-CoV-2 (strain 2019-nCoV/USA-WA1/2020, VR-1986HK from ATCC), hereafter referred to as ATCC-VP. This has been widely used as a quantitative control in molecular assays and to evaluate the efficiency of several workflows, with the advantage of requiring only a BL2 lab [38, 39]. We tested two different numbers of genome copies in a 10 µL volume that we loaded on the metal probe: 2378 and 220000. We calculated these numbers from the original concentration specified by the manufacturer. See “Heat extraction” in “Material and Methods” for details. Before the start of each extraction test, the metal collectors were thoroughly cleaned to eliminate any contamination by RNA or RNase (see “Materials and Methods”). In the biosafety cabinet, we placed one clean metal collector in the center and turned off the air flow. The 10 µL solution was loaded onto the surface of the metal collector in droplets of 1 µL each. We first positioned two droplets symmetrically along the length of the metal collector, and then rotated the it by 72 degrees to repeat this action at a new location until all 10 droplets were loaded. During the loading, the airflow of the biosafety cabinet was turned off and droplets were distributed evenly across the surface of the probe. When loading was completed, the metal collector was left to dry in air for 5 minutes, at the end of which nearly all loaded material had evaporated. At this point, we recollected the metal probe into a 2 mL microcentrifuge tube, added in 220 µL of nuclease-free water, and ensured the entire metal surface was immersed (Fig. 4(a)).

**Figure 4:**
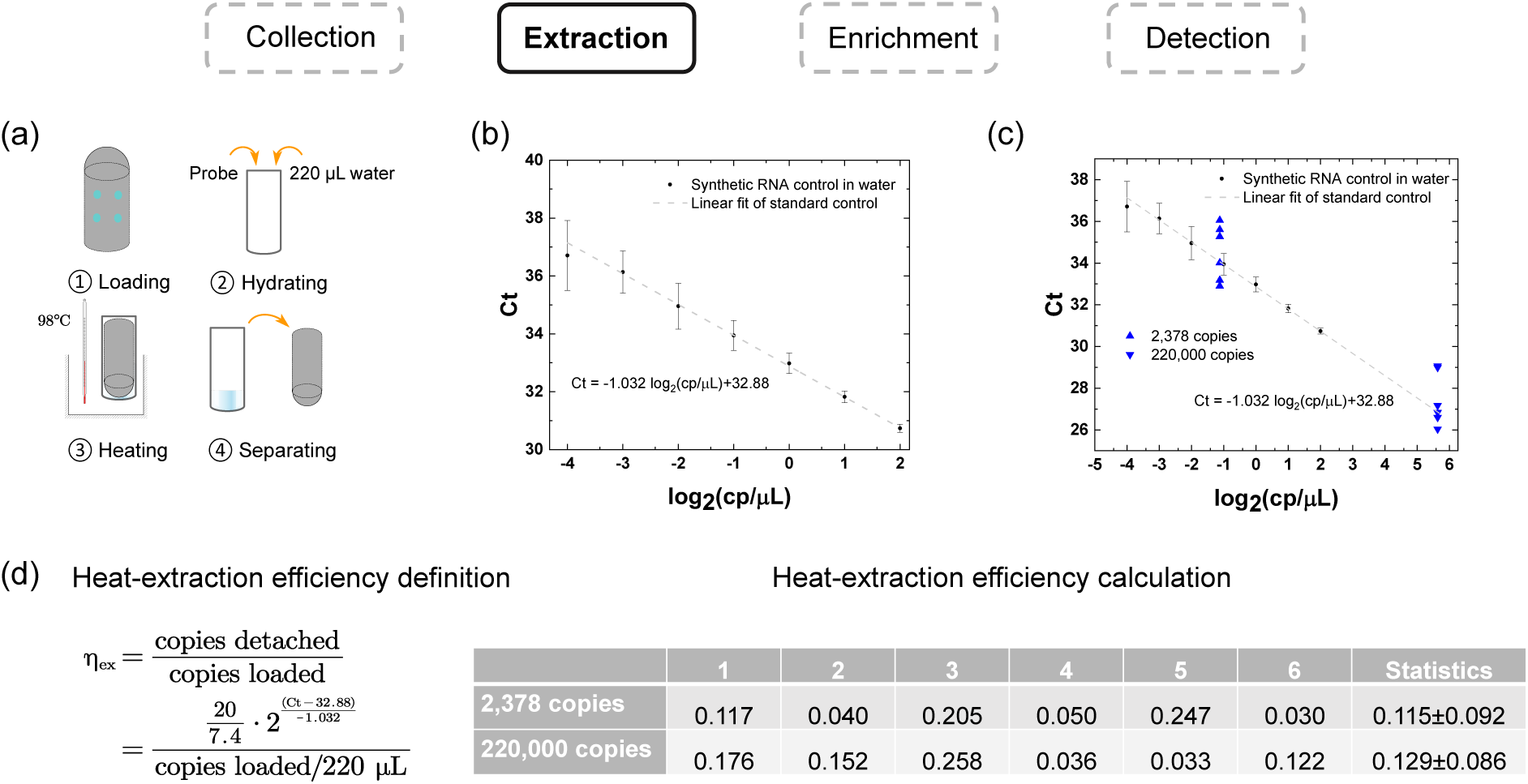
Heat extraction characterization. (a) A fixed number of genome copies of ATCC virus in a 10 µL volume is loaded on the metal probe in 1 µL droplets, dried in air for 5 minutes, and then collected in 2 mL micro-centrifuge tubes. The tube is then heated at 98 °C for 5 minutes. The metal probe is removed after cool down. (b) RT-qPCR standard curves using synthetic SARS-CoV-2 RNA in water (N=24) (see “RT-qPCRs” in Materials and Methods for details). (c) Ct values after heat extraction and RT-qPCR corresponding to the two different numbers of genome copies plotted as blue triangles on the standard curve. For each copy number, N=6 biological replicates were obtained. See Table S1 and S2 for complete data, including negative control (no-heating) and positive control (same number of copies in water). (d) Definition and calculation of the heat-extraction efficiencies with two different genome copies.

To release RNA from the viral particles, we placed the tube containing the metal collector in an OniLAB Mini Dry Bath Incubator preheated to 98 °C and incubated for 5 minutes [40, 37], as illustrated in Figure 4(a). After incubation, the tube was immediately transferred to ice and kept there for 5 minutes. Then, we removed the metal probe from the tube with a magnet, and used a 200 µL pipette to collect residual droplets adhering to the surface of the metal collector to maximize recovery. Typically, about 210 µL could be recovered out of the 220 µL input volume. We thus took 7.4 µL to set up a 20 µL RT-qPCR reaction for SARS-CoV-2 RNA target. We separately obtained standard curves for RT-qPCR using synthetic SARS-CoV2 RNA (Fig. 4(b) and Section “Standard curves and Limit of Detection (LoD) of RT-qPCR”). Linear regression was performed to obtain the function correlating the target’s concentration in the RT-qPCR reaction to the Ct value: Ct = *−*1.032 log_2_(copy */* µL) + 32.88. With this relation, we quantified the extraction efficiency as the ratio of recovered genome copies, calculated from the Ct values of RT-qPCR using the standard curve, and the total number of copies loaded on the metal collector, calculated from dilution of the manufacturer-specified concentration (Fig. 4(d)). We conclude that the efficiencies are 0.115 *±* 0.092 and 0.129 *±* 0.086, for the number of copies loaded on the metal probe at 2378 and 220000, respectively.

The large standard deviation can be attributed to a number of factors, including the composition of the input material, which includes proteins, lipid membranes and other cell lysates that may interfere with RT-qPCR, and the damage of the target during heat inactivation at the manufacturing facility. In fact, this material has been heat-inactivated at 65 °C for 30 minutes, so it is possible that the genetic material may have already been exposed or damaged to different degrees. High variability using this material for isolation of viral RNA was reported before [41], indicating the need for quantitative controls with higher integrity and purity for more accurate characterization of the virus extraction efficiency. Nevertheless, to demonstrate that the variation in data is not caused by our loading experimental protocol, we also loaded the metal collector with Fluospheres (430, same as the #B collected at the lowest concentration in Figure 3(d)) and repeated the above procedure, excluding the heating treatment. We measured the number of recovered Fluospheres using flow cytometry. The results reported in Figure S2 support our claim that the variation in outcomes were due to the starting material and not to our protocol.

### 2.3 Development and characterization of nucleic acid enrichment in solution

We built a custom system to enrich the concentration of nucleic acid in solution with the ultimate goal of increasing the sensitivity of the overall virus detection process. The working principle of the enrichment module is as follows. Negatively charged nucleic acid migrates via electrophoresis in the opposite direction of an applied electric field (toward a positive electrode), but can be blocked by a semipermeable membrane allowing for ion exchange but not escape of large biomolecules, thereby causing nucleic acid to build up at high concentration against the membrane (Fig. 5(a)). The enrichment device consists of a polydimethylsiloxane (PDMS) chip with a circular channel passing completely through the longest dimension of the chip. The channel was designed to accommodate exactly 250 µL of fluid (comparable to the fluid volume from the extraction step) when filled entirely. Both ends of the channel were sealed by semipermeable membranes bonded to the ends of the channels using PDMS (Fig. 5(b)).

**Figure 5:**
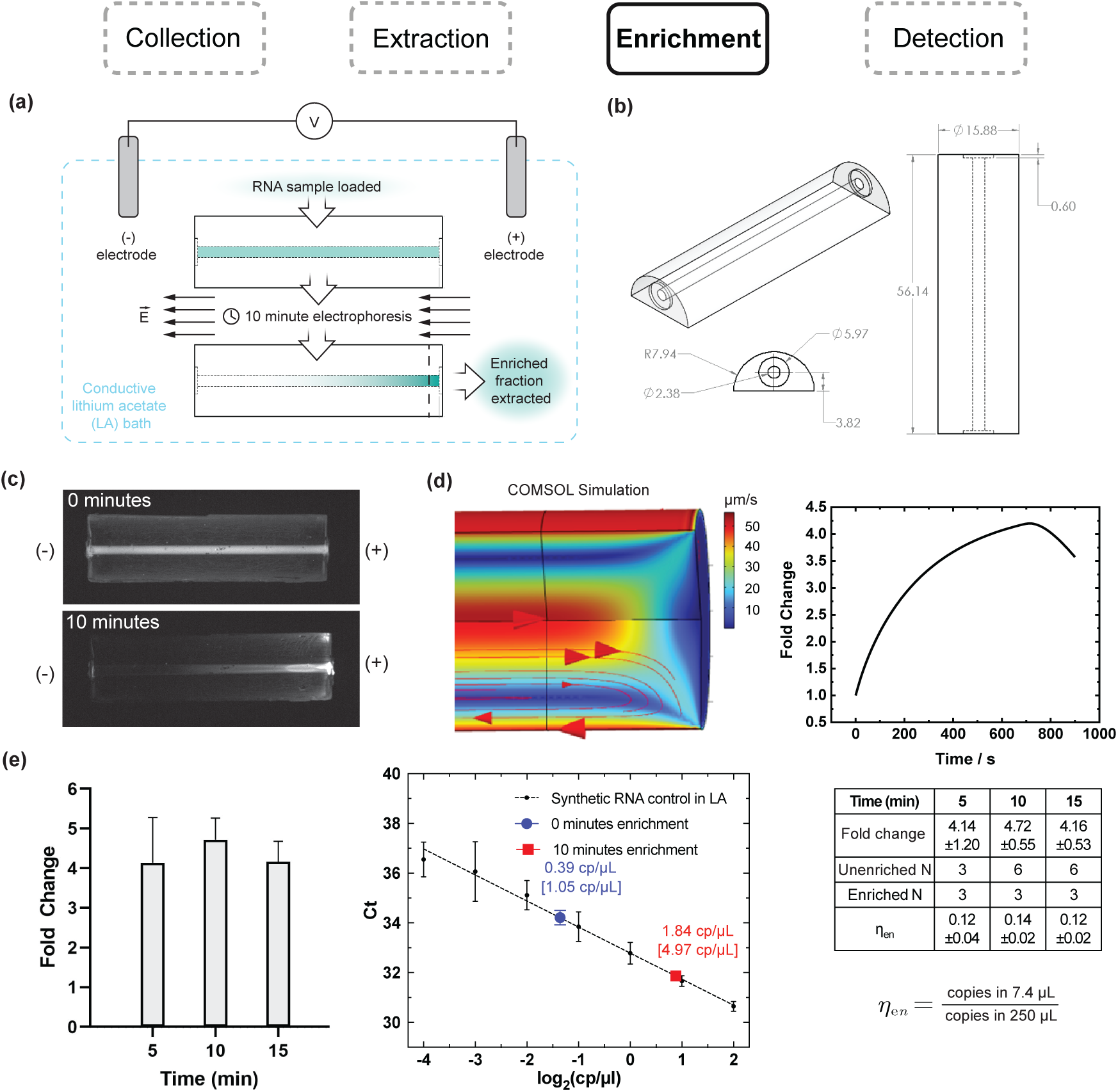
Enrichment of RNA concentration at femtogram to microgram scales through membrane-coupled electrophoresis. (a) Working principle of RNA enrichment. A 250 µL RNA sample buffered in 5 mM lithium acetate (LA) is loaded into a PDMS device consisting of a channel capped at both ends by semipermeable membranes. The device is immersed in an electrophoresis bath containing 5 mM LA and an applied voltage causes electrophoresis of RNA (teal) toward the positive terminal, concentrating the RNA against one of the membranes. An enriched fraction of RNA is then extracted adjacent to the membrane. (b) PDMS channel design for fast enrichment of nucleic acid. (Left) 3D view showing the location of semipermeable membranes in insets. (Center) Face-on view of channel and insets. (Right) Top view, rotated 90 degrees from that shown in (a). Dimensions are indicated in mm. (c) Images of microgram-scale enrichment of mammalian RNA stained with SYBR Green II in devices oriented as in (a), showing concentration profiles before enrichment (0 minutes) and after enrichment (10 minutes) at 2,000 V/m. Both images were obtained using identical exposure settings. (d) Multiphysics simulation of negatively charged particle migration in the enrichment device using COMSOL. (Left) Simulated velocity streamlines. (Right) Simulated time course of concentration fold change adjacent to the membrane (*∼*3% of device volume). For details on the simulations, see “Materials and Methods: Simulation of nucleic acid enrichment in enrichment devices.” (e) Femtogram-scale enrichment of ultra-low concentration synthetic SARS-CoV-2 RNA. (Left) Fold change enrichment of synthetic SARS-CoV-2 RNA quantified by RT-qPCR at 5, 10, and 15 minutes. (Center) 10 minutes of enrichment leads to a 4.7-fold enrichment of RNA. Concentrations before and after enrichment are shown on the RT-qPCR standard curve in blue and red respectively, with bracketed and unbracketed values representing the concentrations in the device and RT-qPCR reactions, respectively. (Right) Summary of data collected, including enrichment efficiency *η*_*en*_.

To perform enrichment, we first determined which conductive medium containing an analyte to be concentrated could be used to fill the channel. We tested the following buffers, including “ultra-fast” electrophoresis buffers: sodium boric acid (10 mM, pH 8), lithium boric acid (1 mM, pH 8.2), lithium acetate (5 mM) [42], and Tris-acetate (40 mM Tris, pH 8.3). We determined their influence on RT-qPCR by detecting known amounts of synthetic viral RNA via RT-qPCR in these buffers. All buffers were compatible with the Luna Universal One-Step RT-qPCR kit (New England BioLabs, E3005), but lithium acetate (LA) demonstrated both desirably low thermal convection due to minimal Joule heating and a fast running speed (data not shown). After verifying that LA buffer did not affect the RT-qPCR standard curve for detecting SARS-CoV-2 (Fig. 6(a)), we selected LA buffer as the conductive medium of choice for nucleic acid enrichment.

**Figure 6:**
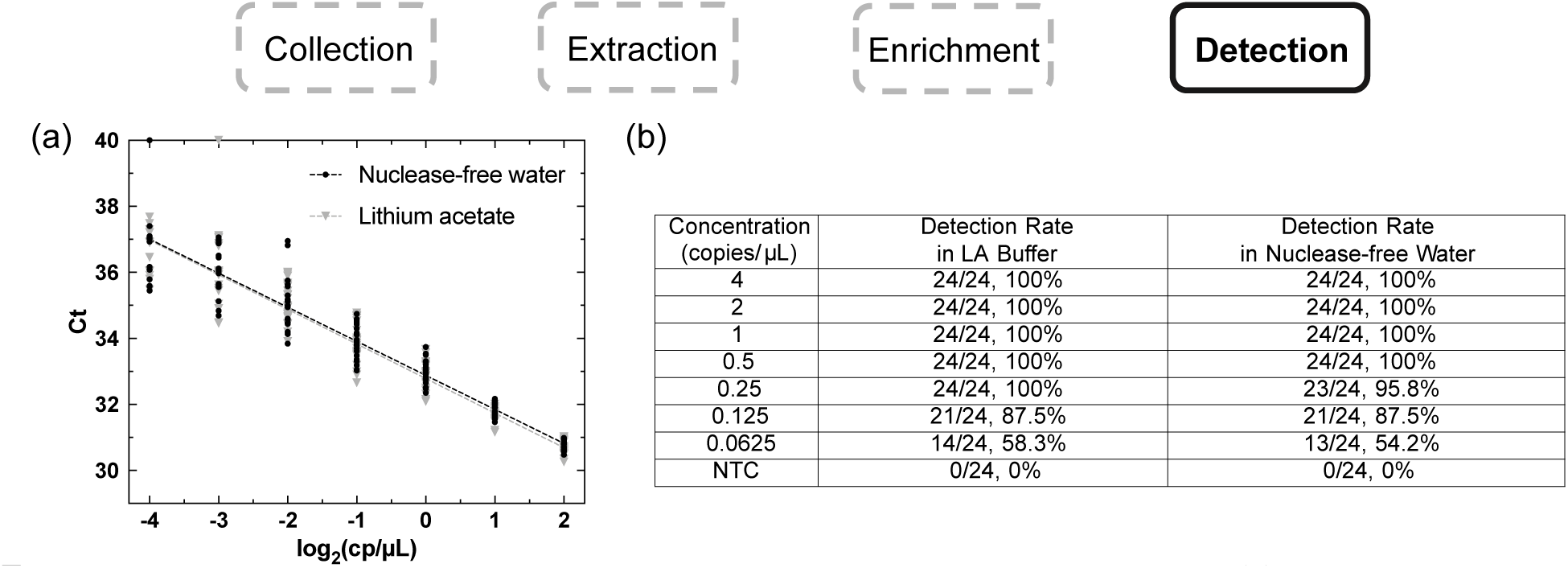
Standard curves of RT-qPCR in nuclease-free water and LA buffer. (a) We performed N=24 replicates for each condition across seven different concentrations using synthetic SARS-CoV-2 RNA. (b) The detection rates at different concentrations for both LA buffer (1.85 mM final concentration in RT-qPCR reaction) and nuclease-free water were reported. The LoDs (limits-of-detection) were determined to be 0.25 copy */* µL for both cases.

After loading the channel with LA buffer containing an analyte, the enrichment module was placed in a standard electrophoresis bath containing LA buffer at 5 mM and a 300 V potential was applied between the electrodes, generating an electric field of approximately 2,000 V/m for a defined duration. See “Methods and Materials: Fabrication of enrichment devices” and “Methods and Materials: Quantification of RNA enrichment” for detailed explanations of the fabrication and enrichment procedures.

To demonstrate the feasibility of the enrichment process, the device was first tested by loading 1 µg of mammalian RNA obtained from HCT 116 cells and stained with SYBR Green II. The imaging results are shown in Fig. 5(c), which demonstrates an evolution from a uniform RNA concentration at 0 minutes to a significant concentration gradient along the channel with enrichment at the positive terminal after only 10 minutes. We noted that the solution concentrates most strongly in the region directly adjacent to the membrane, suggesting that only a small extraction volume is needed to recover a highly concentrated RNA sample. The device was also tested using SYBR Safe™ Stain to visualize DNA (Thermo Scientific™, GeneRuler 1kb DNA Ladder, SM0312) migration over time (Fig. S5). Together, these results confirmed that both DNA and RNA molecules can move toward and concentrate at the membrane adjacent to the positive terminal.

To understand the physical parameters affecting device performance, we performed a multiphysics simulation of the device using COMSOL (Fig. 5(d)). This simulation incorporated electrokinetics and convective fluid flow (refer to “Materials and Methods: Simulation of nucleic acid enrichment in enrichment devices” for simulation details), indicating that the concentration profile adjacent to the membrane is dictated by competing electrophoretic and electroosmotic effects. Electroosmotic flows dominate at longer time scales, causing the concentration of particles near the membrane to reach a maximum and eventually decrease. The simulated maximum degree of enrichment appeared to occur between 10 and 13 minutes.

To explore the required running time and estimate the fold-change after the enrichment, we performed enrichment on ultra-low concentration of synthetic SARS-CoV-2 RNA at 1.2 copies/µL, or 300 copies in 250 µL. The total mass loaded in the device was less than than 4 femtograms. The Ct values obtained before and after enrichment were compared to calculate a fold change in RNA concentration. We found that the maximal fold change was approximately 5-fold after 10 minutes of enrichment (Fig. 5(e)). The maximum degree of enrichment around 10 minutes qualitatively matched the simulation result shown in Fig. 5d. We concluded that use of this enrichment module ought to improve the detection rate of the overall pathogen detection process.

### 2.4 Standard curves and Limit of Detection (LoD) of RT-qPCR

The LoD of RT-qPCR was defined as the lowest concentration of SARS-CoV-2 (copies/µL) at which the detection rate was 95% or higher. In order to determine this concentration, serial dilutions of quantitative synthetic SARS-CoV-2 RNA (ATCC VR-3276SD, lot number: 70048443) at known concentrations were tested for detection using RT-qPCR. For each concentration, 24 replicates of SARS-CoV-2 RNA diluted in either nuclease-free water or LA buffer were tested for detection. Respectively, 24 replicates of non-template controls (NTCs) in either nuclease-free water or LA buffer were also tested. N2 primers [43] were utilized in all RT-qPCR reactions. For both samples diluted in nuclease-free water or LA buffer, the LoD obtained was 0.25 copies/µL (Figure 6). NTCs in either nuclease-free water or LA buffer showed no detection (Figure 6, S6).

We then evaluated the LoD of FAST RT-qPCR with the Chai Open qPCR machine (ChaiBio, single channel) that provides a maximal ramp rate of 5 °C per second. Specifically, we adopted the method of FAST RT-PCR [44] to set up a 20 µL reaction for a selected RNA target. The reaction uses SuperScript IV Reverse Transcriptase (Thermo Fisher Scientific, 18090010) and SpeedStar HS DNA polymerase (TaKaRa, RR070B) to conduct an ultrafast one-step RT-qPCR. As before, the N2 primers were used with the same synthetic SARS-CoV-2 RNA control. The composition of the reaction is listed in Table 4. The thermal cycle comprises the reverse transcription (RT) at 50 °C for 1 minute and the RT deactivation/qPCR activation at 95 °C for 1 minute and the 40 cycles of the denaturation at 95 °C for 1 second and the annealing/extension at 55 °C for 1 second. The Chai Open qPCR machine takes about 24 minutes to finish the defined thermal cycle. The LoD of the FAST RT-qPCR is 0.25 copy */* µL (Figure S4, panel (a), N=32).

### 2.5 Calculation of overall process efficiency

All the efficiencies evaluated are characterized by a mean value, 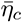, 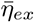 and 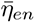, and a standard deviation *σ_c_*, *σ_ex_*, and *σ_en_*, respectively. Specifically, given the evaluations of the efficiencies in Figure 3 (*η_c_* = 0.10*±*0.001),

Figure 4 (*η_ex_* = 0.115 *±* 0.09) and Figure 5 (*η_en_* = 0.14 *±* 0.02), the mean value 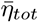 is given by

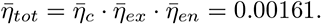

The standard variation *σ_tot_* of the whole process efficiency can then be evaluated by exploiting the formula for the uncertainty propagation of a product, that is,

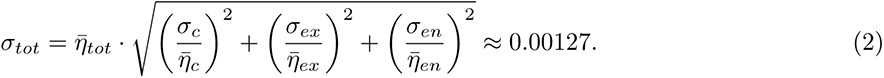

From this, the mean concentration in air that we can detect is 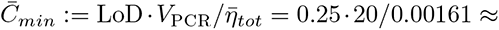 3106 particles/*m*^3^. From this calculation, it is possible to conclude that the minimal concentrations that our device is able to detect are roughly between 649 and 5562 particles/*m*^3^. These fall in the lower part of the concentration range of influenza virus detected in enclosed environments during flu season [36] or of SARS-CoV-2 virus detected in hospital rooms [17].

## 3 Process integration and detection rate evaluation

To evaluate the sensitivity and specificity of our system, from Collection to Detection, we again employed heat-inactivated SARS-CoV-2 viral particles as the starting material to carry out the entire process. All metal probes were thoroughly cleaned following the protocol described in “Metal collector cleaning” in “Materials and Methods”.

For each test, as shown in Figure 7, we started with a series of 10-fold dilutions of ATCC-VP and aliquoted 10 µL at the end containing a defined number of copies of ATCC-VP. We experimented with two different numbers: 430 and 500 copies. The first number corresponds to the number collected on the metal probe in correspondence to the lowest beads concentration in air tested (Figure 3(d)). We also chose 500 copies to explore the limit of detection. Before loading the ATCC-VP onto the metal collector, all working surfaces and equipment were wiped with RNaseZap™ Solution, and the ventilation of the biosafety hood was turned off to minimize evaporation during loading. A clean metal collector was picked from the sealed container using a tweezer and placed upright on a magnet. A 2.5 µL pipette was used to transfer 1 µL out of the aliquoted 10 µL solution onto the surface of the metal probe (see “Heat Extraction” in “Materials and Methods” for how droplets were loaded onto the probe). After loading, the metal probe was left to dry in the biosafety cabinet for 5 minutes as before (ventilation off). The probe was then collected in a 2 mL microcentrifuge tube with 220 µL of nuclease-free water. The tube and metal probe were heated in the bath at 98 °C for 5 minutes before being transferred to ice to cool down. The tube was then brought back to the biosafety hood and the metal collector was removed with a magnet. We then measured the volume of the liquid left in the 2 mL tube. The number was later used to calculate how much additional buffer was needed to fill the enrichment device channel.

**Figure 7:**
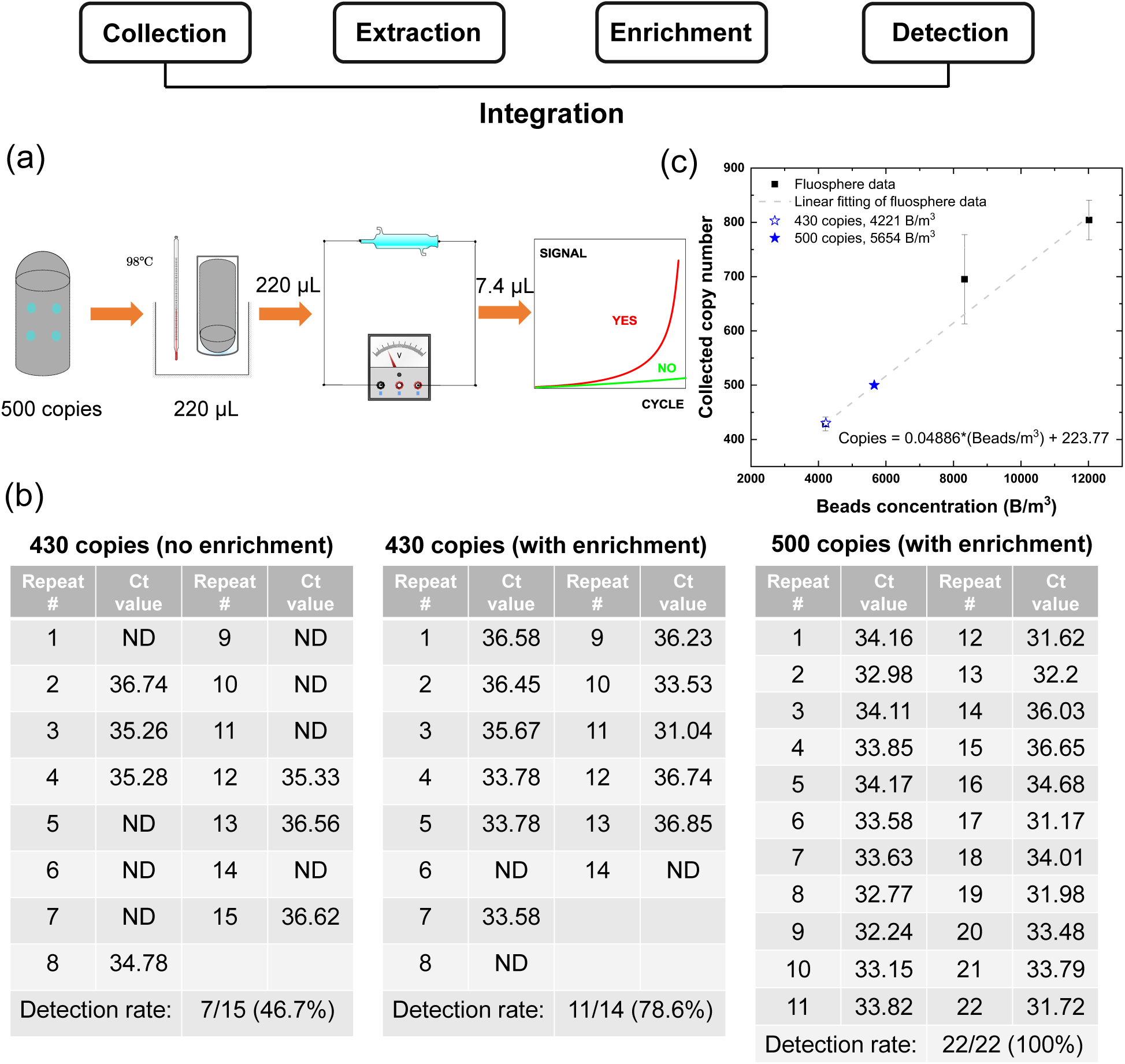
Experimental process for the integration test. (a) **Loading:** the metal probe was loaded with either nuclease-free water (for NTCs) or SARS-CoV-2 ATCC-VP (for biological replicates) using 1 µL droplets with a total volume of 10 µL. **Extraction:** the metal probe was dried in a biosafety cabinet, collected in 2 mL tubes and 220 µL of water was added before heating at 98 °C for 5 minutes. **Enrichment:** the recovered liquid was supplemented with LA buffer and concentrated in the enrichment device for 10 minutes. **Detection:** 7.4 µL of enriched liquid was extracted from the positive end of the enrichment device and analyzed with RT-qPCR to determine the Ct value. (b) Results of integration test. The starting copies and enrichment conditions are specified at the top. NTCs were all negative and can be found in the “Integration” of Supplementary Materials (Table S3).

For the 430 copy experiment, after the heat-extraction, we followed two parallel routes to evaluate the benefit of incorporating the enrichment process for detection. Specifically, in the first case, we aliquoted 7.4 µL from the recovered solution (averaged 210 µL) and made a 20 µL RT-qPCR reaction to analyze (no enrichment). The rest of the solution was supplemented with 13 µL of PCR-grade LA buffer stock (20x) and nuclease-free water to make a total input volume of 250 µL to the enrichment process (Figure S3). Because of the extraction of this 7.4 µL, we effectively removed 15 copies, leaving us with 415 copies. Also, due to a loss of 0.5% of the volume during loading 1 µL droplets, we eventually reached an equivalent starting number of 394 particles, which implies that we are underestimating our performance (see “Integration” in Supplementary Materials). This mixture was thus injected into the enrichment device and the process described in Section 2.3 was followed, running three devices at a time. At the end of enrichment, exactly 7.4 µL of sample was recovered near the anode of each device with a 10 µL pipette as before, followed by addition of RT-qPCR reagents to make a 20 µL reaction volume. The Ct values of all the replicates are reported in Figure 7(b), left and middle tables. Quantification of detection before and after enrichment at 430 copies shows that enrichment boosted the detection rate from 46.7% to 78.6% for N=15 and 14, respectively. For the 500 copy experiment, we only performed the process containing enrichment and obtained a 100% detection rate for N=22 (Figure 7(b), right table).

For the non-template controls, we also loaded the metal probe with 10 µL of nuclease-free water and went through the same process. Again, we aliquoted 7.4 µL for RT-qPCR analysis before enrichment and filled the rest of solution with 12.5 µL of 20x LA buffer, injecting a 250 µL volume into the enrichment device to obtain data after enrichment. None of the NTCs were amplified (0/15) (top left (before enrichment) and right (after enrichment) of Table S3), demonstrating the specificity of the process.

To convert the copy numbers loaded on the metal probe to equivalent particle concentrations in air, we fitted the three data points corresponding to the lowest concentrations in Figure 3(d) with a line. We then plotted the two copy numbers (430 and 500) against the fitted line and calculated the corresponding bead concentration, as shown in Figure 7(c). Overall, we estimated the beads per m^3^ corresponding to the 500 loaded copies as *C_min_* = 5654 B*/*m^3^. Given a 100% detection rate, this is an upper bound on our limit of detection as determined by the integration test. For the copy number 430, corresponding to 4221 B/m^3^, we achieved a 46.7% detection rate without enrichment and a 78.6% detection rate with enrichment. So, this concentration is a lower bound on our limit of detection as determined by the integration test.

## 4 Conclusions and Discussion

In this paper, we have characterized the performance of a rapid fieldable airborne virus detection process that includes an enrichment step to concentrate RNA in a small volume. Specifically, our enrichment system uses capillary electrophoresis against a semipermeable membrane, and is able to concentrate RNA by about 5-fold. By virtue of this enrichment system, we could reach a sensitivity of about 5,000 copies of virus per m^3^ of air, a concentration that falls in the lower range of influenza virus and SARS-CoV-2 concentrations reported in the literature [45, 17]. By including our enrichment system in the detection process, we therefore make airborne virus detection sufficiently sensitive for practical application. Being based on RT-qPCR, the process that we have proposed is applicable to any other virus whose genetic sequence is known. At the same time, our enrichment process can be used in a number of other applications including nucleic acid fragment analysis and quality control, next-generation sequencing, and general biomolecular analysis. Here, we designed the enrichment device to be simple and fieldable, but incorporation of parallelized microcapillaries, fluidic manifolds [46], electric field gradient focusing, or isotachophoresis [47, 48] could further improve the degree of enrichment at the expense of additional cost and complexity.

Although we utilized standard RT-qPCR reagents, lyophylized reagents are appearing [49]. These would allow us to remove the three-fold dilution factor when taking the 7.4 uL to the 20 uL volume PCR reaction and hence could decrease the sensitivity by up to a factor of three, bringing the sensitivity to *C_min_≈* 2, 000 cp/m^3^. However, more work needs to be done in order to determine the extent to which with lyophilized reagents the same RT-qPCR LoD can be maintained in LA buffer and with the fast RT-qPCR.

There are also several sources of error in our quantification process that could be responsible for under-estimating the system’s performance. Specifically, the use of the ATCC inactivated virus is one such source of error. In fact, being heat-inactivated, many of the RNA particles are likely already exposed and can be easily damaged during the heat extraction process. This is consistent with our finding that when performing heat extraction in water, we achieve lower output than just analyzing the unheated water with RT-qPCR (Tables S1, S2). Another such source of error is the treatment of a fluorescent bead as if it were just one viral particle, while in practice solid aerosol of that size may include multiple virus genome copies [50, 51].

Future studies will be carried in the field, with positive and negative control rooms, in order to confirm selectivity and sensitivity in a real-world setting.

## Materials and Methods

### Inactivated viral particles

Heat-inactivated SARS-CoV-2 (strain 2019-nCoV/USA-WA1/2020, VR-1986HK) was obtained from ATCC. Lot number:70037781. This material has been inactivated by heating at 65 °C for 30 minutes, and we aliquoted the original stock upon receipt to minimize the number of freeze-thaw cycles and ensure consistency by using one of the same batch of aliquoted samples for each experiment.

### Synthetic SARS-CoV-2 RNA

Quantitative synthetic SARS-CoV-2 RNA was obtained from ATCC (VR-3276SD, lot number: 70048443) and used as standard controls for characterization of the RT-qPCR assay, and evaluation of the performance of the electrophoresis enrichment system. The synthetic RNA contains fragments from the ORF1ab, E, and N regions of the SARS-CoV-2 genome.

### RT-qPCRs

The RT-qPCR reactions were prepared inside a PCR-dedicated laminar flow hood. The hood was sterilized by wiping the surface with PREempt wipes and irradiating with UV light for 30 minutes. All lab work was conducted while wearing a N-94 mask, and materials and reagents were opened only inside the hood. A clean-to-dirty workflow was followed, where we first prepared the RT-qPCR reactions by mixing the Luna Universal One-Step Reaction Mix, Luna WarmStart RT Enzyme Mix, and primers, while keeping the SARS-CoV-2 RNA sealed and outside of the hood. We then brought the SARS-CoV-2 RNA into the hood, performed serial dilutions (either in nuclease-free water or lithium acetate), and mixed the RNA with the rest of the reagents to prepare the final RT-qPCR reactions. For reactions in LA buffer, the final concentration of LA was 1.85 mM. The Luna Universal One-Step RT-qPCR kit (NEB, E3005) was utilized for all RT-qPCRs in a Roche LightCycler 480. For each RT-qPCR reaction, the SARS-CoV-2 sample was suspended in a total volume of 7.4 µL in either nuclease-free water or LA buffer (5 mM). Subsequently, the 7.4 µL sample of SARS-CoV-2 was added to make a final volume of 20 µL according to Table 2 (1.85 mM final concentration of LA). The timing and temperatures utilized are shown on Table 3. N2 forward (IDT, 10006824, 5’-TTACAAACATTGGCCGCAAA-3’) and N2 reverse (IDT, 10006825, 5’-GCGCGACATTCCGAAGAA-3’) primers were utilized in all reactions. To verify that the reagents, materials, and preparation zone remained uncontaminated in each experiment, 12 NTC wells were prepared on every RT-qPCR plate used in the study and verified to be negative after 45 cycles.

**Table 1:** Comparison of the detection methods in terms of LoD and running time. Synthetic SARS-CoV-2 RNA in LA buffer is the target to be detected with the indicated primers (and probe for Fast RT-qPCR).

| Method | LoD in LA buffer (copy/ $\mu\text{L}$) | Running Time (minute) | PCR machine |
| --- | --- | --- | --- |
| RT-qPCR | 0.25 | $\sim 58$ | Roche LightCycler 480 |
| Fast RT-qPCR | 0.25 | $\sim 24$ | Chai Bio Open PCR |

**Table 2:** RT-qPCR reaction composition for SARS-CoV-2 detection in nuclease-free water or LA buffer

| Component | 20 μL reaction | Final concentration |
| --- | --- | --- |
| Luna Universal One-Step Reaction Mix (2X) | 10 μL | 1X |
| Luna WarmStart RT Enzyme Mix (20X) | 1 μL | 1X |
| N2 Forward Primer (10 μM) | 0.8 μL | 0.4 μM |
| N2 Reverse Primer (10 μM) | 0.8 μL | 0.4 μM |
| SARS-CoV-2 RNA in water or LA buffer | 7.4 μL | variable |

**Table 3:**
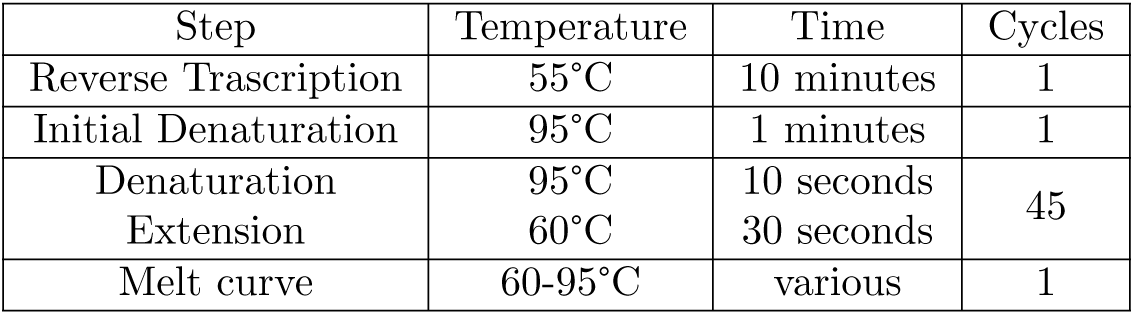
RT-qPCR timing and temperatures.

**Table 4:** Composition of Fast RT-qPCR reaction. *^a^*, The forward/reverse primers and the probe should be sequence-specific to the target template and are dissolved in nuclease-free water. Here, we target for the N gene of SARS-CoV-2 RNA genome. *^b^*, Synthetic RNA or analyte can be in nuclease-free water or LA buffer. *^c^*, Nuclease-free water should be replaced with 1x LA buffer when synthetic RNA or analyte is in LA buffer.

| Stock solution | Supplier | Final concentration | unit | 20 μL reaction (μL) |
| --- | --- | --- | --- | --- |
| 10 mM dNTPs | New England Biolabs (N0447S) | 0.12 | mM | 0.24 |
| 5x SuperScript IV Reverse Transcriptase buffer | Thermo Fisher Scientific (18090010) | 0.3 | x | 1.2 |
| 100 mM DTT | Thermo Fisher Scientific (18090010) | 1 | mM | 0.2 |
| SuperScript IV Reverse Transcriptase (200 U/μL) | Thermo Fisher Scientific (18090010) | 2 | U/μL | 0.2 |
| SpeedStar HS DNA polymerase (5 U/μL) | TaKaRa (RR070B) | 0.04 | U/μL | 0.16 |
| Fast Buffer I (10X) | TaKaRa (RR070B) | 1 | x | 2 |
| N2 forward primer (10 μM)<sup>a</sup> | Integrated DNA Technology | 0.4 | μM | 0.8 |
| N2 reverse primer (10 μM) | Integrated DNA Technology | 0.4 | μM | 0.8 |
| N2 probe (10 μM) | Integrated DNA Technology | 0.2 | μM | 0.4 |
| synthetic RNA (10x)<sup>b</sup> | Codex (SC2-RNAC-0100) | 1 | x | 2 |
| *nuclease – freewater*<sup>c</sup> | Zymo research (W1001) | – | – | 12 |

### Use of counting beads for flow cytometry measurements

To ensure more reliable results in our collection experiments, we added CountBright Absolute Counting Beads (CBs) in our samples before flow cytometry analysis. CountBright Absolute Counting Beads are a calibrated suspension of brightly fluorescent microspheres, containing a known concentration of microspheres. To accurately determine the absolute particle counts in our samples, a specific volume of the CBs was added to the samples. This allowed us to analyze a fraction of the sample volume and precisely determine the total number of particles in our samples using the following formula:

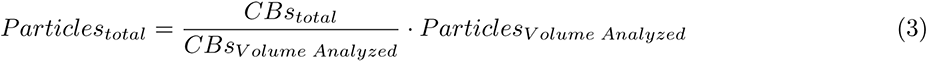

### Metal collector cleaning (0.5% hydrogen peroxide)

The metal collectors were manufactuered at machine shops. CNC coolant or oily residues on the surface of the metal probe could interfere with the electrostatic precipitation and negatively affect the performance of the collection. In order to optimize the process, we placed the metal collectors in the sonicator with MiliQ water, and then ran the sonicator for 20 minutes. We then treated the surface of the metal collectors with the PREempt RTU disinfectant solution that contains 0.5% hydrogen peroxide by soaking overnight. Prior to each Collection test, we used a magnet and a PREempt Wipe to take a clean probe out of PREempt Solution, with no contact with the metal piece directly. Then we used the same wipe to rub the entire surface of that metal piece thoroughly (making sure that the PREempt solution residue is uniform on the surface), then put it on ESP device. We sealed the tent and let the metal collector to dry in air for 5 minutes before plugging in the nebulizers and ESP device at the same time to inititate collection.

### Metal collector cleaning (molecular biology grade)

Before each integration and heat-extraction test, each metal collector was immersed in 1% Alconox™ Detojet™ Low-Foaming Liquid Detergent for overnight. This ensures that any residual RNA would be chemically degraded [52]. After that, the metal collectors were rinsed thoroughly in nuclease-free water for three times until no visible foams can be seen. Then, we used the RNaseZap™ RNase Decontamination Solution to spray over the metal collectors to keep in contact for 5 minutes, before thoroughly rinsed in nuclease-free water for another three times until no visible bubbles can be seen. This prevented any potential RNase contamination. The metal collectors were then autoclaved using the sterilize + dry cycle to further keep them dry and clean. The metal collectors’ container was then sealed and opened only under biosafety cabinet.

### Heat extraction

To prepare solutions containing a specific number of genome copies of viral particles, we performed a series of 10-fold dilution of the Heat-inactivated SARS-CoV-2. As the stock contains cell lysate and supernatant from Vero E6 cells which were used by the manufacturer to propagate the viruses, special care should be taken to sufficiently mix at each step of dilution by pipetting up and down over 50 times in low-binding tubes. At the end of dilution, we aliquoted 10 µL of solution containing targeted numbers of genome copies of the virus as calculated from the original concentration specified by manufacturer. The aliquoting of 10 µL ensured we were loading the same number of genome copies across all biological replicates and improved our consistency (see Figure S2). The two targeted numbers of copies correspond to the the maximum concentration tested in generating RT-qPCR standard curves (see Figure 6(a)), and a concentration of 1000 copies */* µL when resuspended in 220 µL of water. During loading, we used a 2.5 µL pipette to transfer the first droplet of 1 µL to approximately 1/3 of the length from the top of the metal collector, and a second droplet was placed at approximately 1/3 from the bottom. Then, the metal collector was rotated about 72 degrees and repeated this process, until all 10 droplets were distributed evenly over the surface. Since we used a new pipette tip for every 1 µL droplet, 0.5 µL of the 10 µL was lost because small amount of liquid adhered to the inside of pipette tips. We adjusted the genome copy numbers calculation to account for this fact. All the loading the drying were performed in a biosafety cabinet with ventilation turned off. After heating, each tube was immediately placed in ice to cool down. We took a 7.4 µL heated sample to set up a 20 µL RT-qPCR reaction.

### Fabrication of enrichment devices

Enrichment devices were fabricated by casting PDMS (Sylgard 184, Fisher Scientific, NC9285739) in custom-designed molds. The molds were created using a two-step process. First, the body of each mold was designed in SolidWorks and 3D-printed using a Formlabs stereolithography printer with clear resin (purchased through Protolabs). The geometry of the mold consisted of a semicircular sink with two 2.38 mm diameter holes bored through the flat ends (Fig. 5a). After printing, molds were washed with 100 % isoproanol and cured in a UV oven for 30 minutes. Next, 7.5 cm stainless steel rods of 2.38 mm diameter were lightly greased with commercial petroleum jelly and inserted through the holes in each mold to form the channel running axially through the device (Fig. 5a). Plastic spacers were used to create a press fit between the rod and the holes in the 3D printed part. The PDMS elastomer was prepared according to the manufacturer’s instructions, mixed by vortexing, then cast in the molds. The PDMS-containing molds were degassed for 10 minutes and baked in a 65 °C oven for 2 hours. Excess petroleum jelly was removed from the channel walls using a clean room swab. The rod was extracted and the PDMS cast was removed using forceps. The first PDMS cast created in each mold was discarded.

After creation of the PDMS devices shown in Fig. 5a, semipermeable membranes were attached to either end of the channel using PDMS as an adhesive. Briefly, double-layer cellulose 12-14 kDa molecular weight cutoff dialysis membranes (Spectrum Labs, 132678T) were cut into circles of 5 mm diameter using a biopsy punch (World Precision Instruments, 504532). Degassed uncured PDMS was applied to the flat end of the PDMS cast making a circle around each hole using a syringe with a blunt-end needle as an applicator. The applied PDMS was smoothed to a uniform thickness using a micropipette tip. Using forceps, the double-layer semipermeable membrane cutouts were centered on each hole and gently applied to the PDMS to create a seal. The PDMS sealant was allowed to cure in a 65 °C oven for 60 minutes. Then, a second ring of PDMS was applied to the outer portion of each membrane to improve the quality of the seal. The PDMS was cured at 65 °C for an additional 60 minutes.

### RNA enrichment and quantification

To test the performance of the enrichment device, synthetic SARS-CoV-2 RNA (ATCC, VR-3276SD) was diluted to a target concentration of 1.2 copies/µL in 250 µL a 5 mM lithium acetate (LA) buffer optimized for fast electrophoresis and minimal Joule heating [42]. After addition of the RNA to the buffer, the sample was homogenized by pipetting. Before loading the PDMS devices, a portion of the sample was set aside on ice for RT-qPCR analysis. Devices were loaded with sample by injection using a syringe. Briefly, two 23 gauge needles (BD, 305194) were pierced through the PDMS adjacent to each membrane. One needle was used to fill the channel by slow injection of a 250 µL sample from a syringe, while the other needle served as an air escape. Following injection of the sample, any bubbles created were collected near the air escape needle and removed with a syringe, allowing fluid from the sample injection needle to replace the volume. The needles were then removed, leaving no visible holes in the PDMS.

After loading, the devices were placed in a standard benchtop gel electrophoresis bath (VWR, 89032-290) with a 15.25 cm distance between electrodes filled with 5 mM LA running buffer. Devices were aligned to place the axis of the channel parallel to the direction of the electric field (Fig. 5a). All tests were conducted using a voltage of 300 V (voltage supply dimensions = 200 x 290 x 70 mm (W x D x H), weight = 1.2 kg), corresponding to an approximate electric field strength of 2,000 V/m or 20 V/cm. Enrichment was allowed to proceed for 5, 10, or 15 minutes.

Upon completion of electrophoresis, devices were gently removed from the running bath and a sample of 7.4 µL was extracted using the following procedure. A 23-gauge needle was first inserted into the channel near the negative-terminal membrane, to provide a fluid escape. To separate the most concentrated 7.4 µL from the remaining 242.6 µL of fluid, a small air gap was inserted with a syringe into the channel at a distance of 1.6 mm from the positive-terminal membrane. The channel was then cut with a razor blade across the air gap, and the 7.4 µL sample was removed using a micropipette and analyzed by RT-qPCR to quantify the concentration of SARS-CoV-2 RNA relative to unenriched control samples.

### Visualization of nucleic acid enrichment in enrichment devices

HCT 116 human colorectal cancer cells (ATCC, CCL-247) were cultured in DMEM (Gibco, 11965084) with 10% fetal bovine serum (Sigma Aldrich, F2442) and 1% penicillin-streptomycin (Life Technologies, 15140122). Total RNA was isolated from these cells using a PureLink RNA Mini Kit (Invitrogen, 12183018A). 1 µg of RNA was diluted in 250 µL of LA to fill each enrichment device. SYBR Green II (Invitrogen, S7564), an RNA-specific nucelic acid stain, was prepared at 1x in the RNA-containing buffer and mixed prior to loading. Devices were imaged on a 470 nm LED transilluminator (IO Rodeo, IMG-04-02) before and after conducting enrichment at 300 V for 10 minutes using identical zoom, illumination, and exposure settings.

To visualize DNA enrichment, SYBR Safe™ Stain was prepared at 1x in LA with 1 µg DNA (Thermo Scientific, GeneRuler 1kb DNA Ladder, SM0312) and imaged every 6 minutes for 30 minutes in the running conditions described above. The device was also loaded with TriTrack three-color DNA loading dye (Thermo Scientific, R1161) alone and imaged after 15 minutes to confirm enrichment visually.

### Simulation of nucleic acid enrichment in enrichment devices

The fluid dynamics simulation was performed using COMSOL Multiphysics. An axisymmetric geometry was used with matching dimensions as specified in Figure 5(b). At the ends of the channel, we applied an voltage of 300 V and water was assumed to be able to flow through the channel opening at a small but constant averaged velocity of 0.01 µm*/* sec to emulate the semi-permeable membranes at two ends. We first simulated the steady flow generated by electroosmosis, and the *ζ*-potential of the PDMS channel was determined to be *ζ*_PDMS_ = *−*5 *∼ −*60mV [53, 54]. The diffusion coefficient of SARS-CoV-2 RNA can be estimated from the molecular dynamics simulations of single-stranded RNA, and it was determined to be 0.68 *∼* 1.13 *×* 10*^−^*^12^m^2^ */* s [55, 56, 57]. And the effective charge was estimated to be *−*64 *∼ −*434 for a 30kb genome [58]. These electrokinetic constants were varied within the above ranges to minimize the difference in outcomes between simulation and experiments. We chose *ζ*_PDMS_ = *−*13 mV, the effective charge *q*_RNA_ = *−*120 and a diffusion coefficient of *D*_RNA_ = 1.13 *×* 10*^−^*^12^m^2^ */* s.

### Fast RT-qPCR composition

## Data Availability

All data produced in the present study are available upon reasonable request to the authors

## Acknowledgements

The authors would like to acknowledge Mehdi Salek, Linda Griffith, Kurt Broderick, and the MIT NEET Living Machines program for assistance with the development of the enrichment module, Dan Gilbert at the MIT Laboratory for Manufacturing and Productivity for assistance with device fabrication, and the MIT BioMicro Center and the labs of Richard Young and Ron Weiss for assistance with RT-qPCR. The authors thank the lab of Nicholas Fang for the help with the COMSOL software simulations. The authors thank Krishna Manoj, Ted Grunberg, and Miranda Cai for assistance with early prototypes and helpful discussions. We thank the Deshpande Center for their support and the AFOSR (grant number FA9550-20-1-0044) for funding.

## 5 Supplementary Material

### Collection

**Figure S1:**
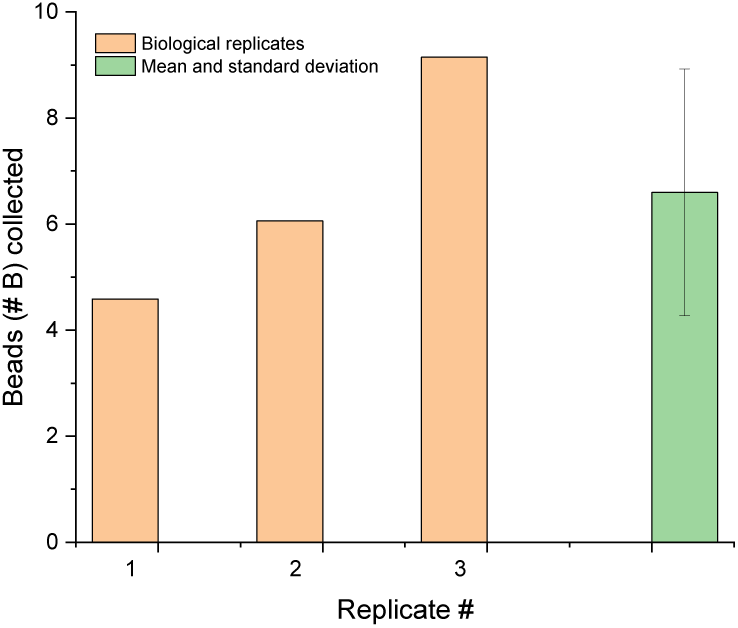
Fluosphere background measurement. We performed three replicates of the background measurement in the Collection experiments, where we nebulized DI water without any Fluosphere added for the same 22 minutes. The metal collectors were treated in the same way as described in Collection.

### Heat-extraction

**Figure S2:**
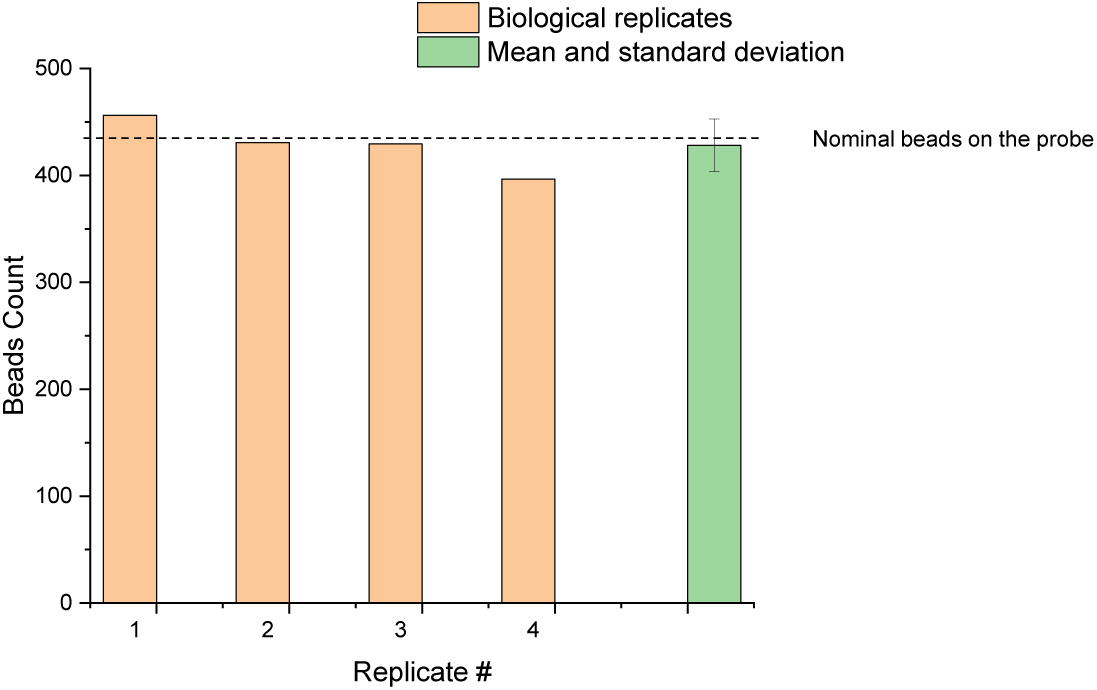
Fluosphere detachment test. To demonstrate the consistency of our loading-extraction protocol, we loaded the metal collectors with the same number of Fluosphere (430, as the lowest concentration of extraction test) and followed the steps outlined in Figure 4(a). Green bar on the right shows the mean and standard deviation over four biological replicates: 428 *±* 24.

**Table S1:**
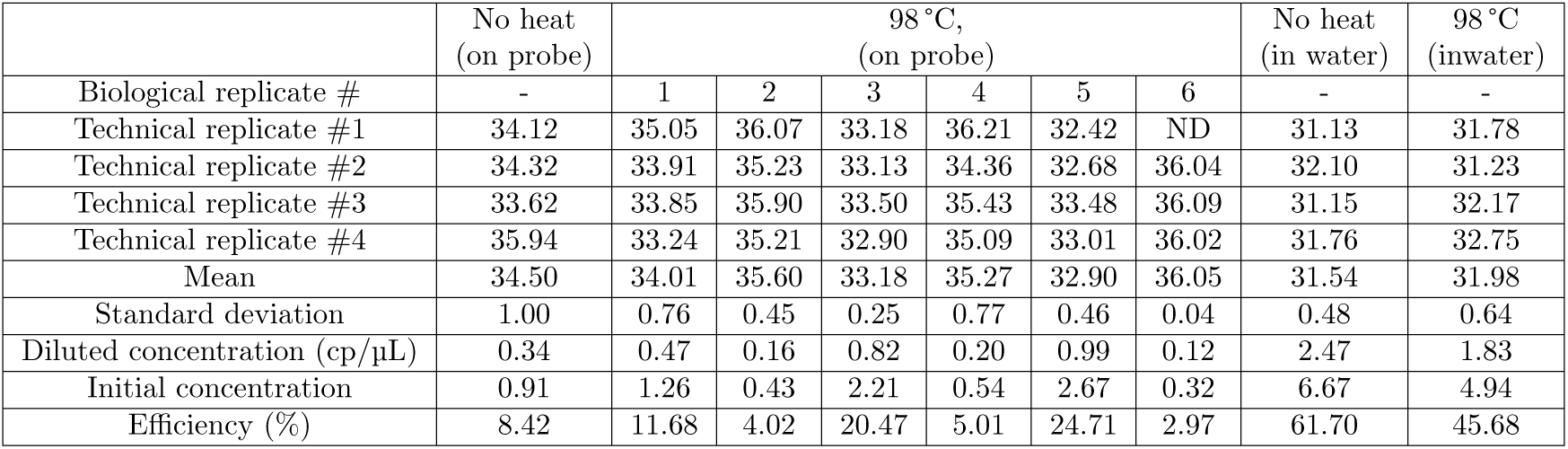
Results of of heat-extraction tests (2378 copies). For each biological/technical replicate, the Ct value was reported. The first column was loading the same number of copies on the probe but with no heating treatment, followed by six biological replicates with heat treatment at 98 °C. Last two columns reported the results of spiking the same amount of copies in water with and without heating treatment.

**Table S2:**
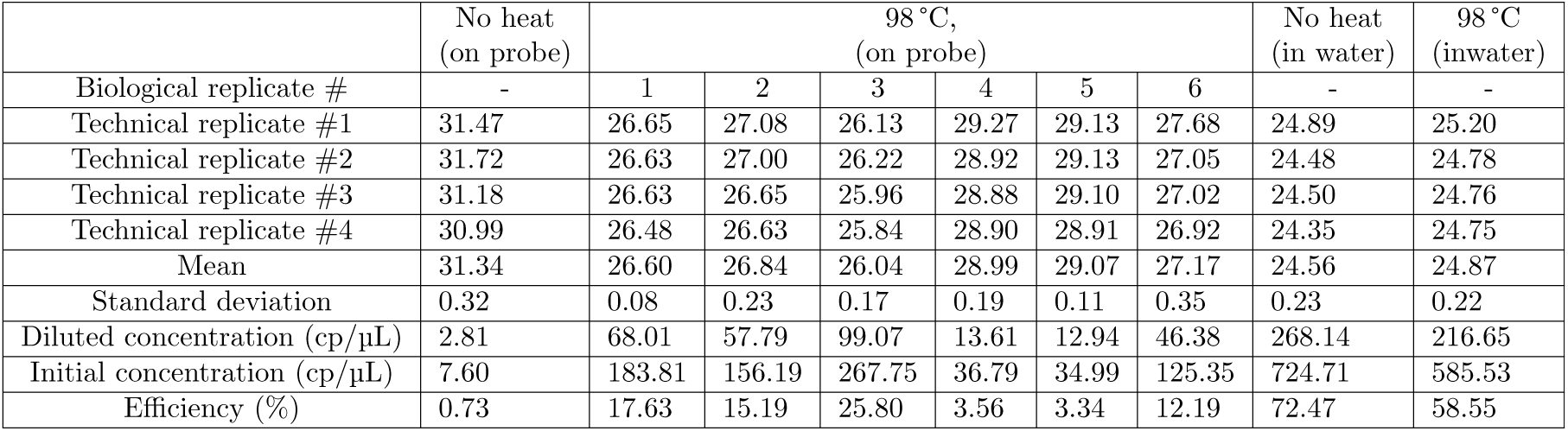
Results of of heat-extraction tests (220000 copies). For each biological/technical replicate, the Ct value was reported. The first column was loading the same number of copies on the probe but with no heating treatment, followed by six biological replicates with heat treatment at 98 °C. Last two columns reported the results of spiking the same amount of copies in water with and without heating treatment.

### Integration

As mentioned in the Integration section of main text, we experimented with two different number of starting genome copies on the metal probe (430 and 500). The first number 430 corresponds to the # B we collected at the lowest concentration in Figure 3(d). From the master stock of ATCC-VP, we performed a series of 10-fold dilutions and made a 10 µL solution containing 430 copies of ATCC-VP. Then we used a 2.5 µL pipette to transfer 1 µL of this solution onto the surface of the metal probe. Due to the loss of liquid (residuals on the inner surface of pipette tips), 9.5 µL was actually loaded. In addition, at the end of heating heat-extraction, usually around 210 µL of liquid could be recovered. We aliquoted another 7.4 µL for RT-qPCR analysis as a control for no-enrichment, and some target molecules were also lost in this process. Eventually, the number of genome copies should be back-calculated as: 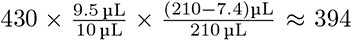 The aliquoting step was also illustrated in the diagram Figure S3. For the 500 genome copies, we did not have any loss due to the pipetting or aliquoting, so corrections were not made for this case.

We also conducted the integration tests for non-template controls (NTCs), and the results were displayed in Table S3.

### Fast RT-qPCR

For Fast RT-qPCR, positive identification rate is 100% (N=32) to detect down to 0.25 copy/µL in LA buffer (Fig. S4(a)). The amplification curves of 0.25 copy/µL in 32 replicates (Fig. S4(b)) clearly shows amplification between 36 and 40 cycles, comparing to the ones of non-template control (i.e., nuclease-free water in 32 replicates (Fig. S4(c)) in LA buffer. This suggested that fast RT-PCR in LA buffer can only answer presence or absence of the synthetic SRAS-CoV-2 RNA analyte but cannot distinguish the amount of analyte.

**Figure S3:**
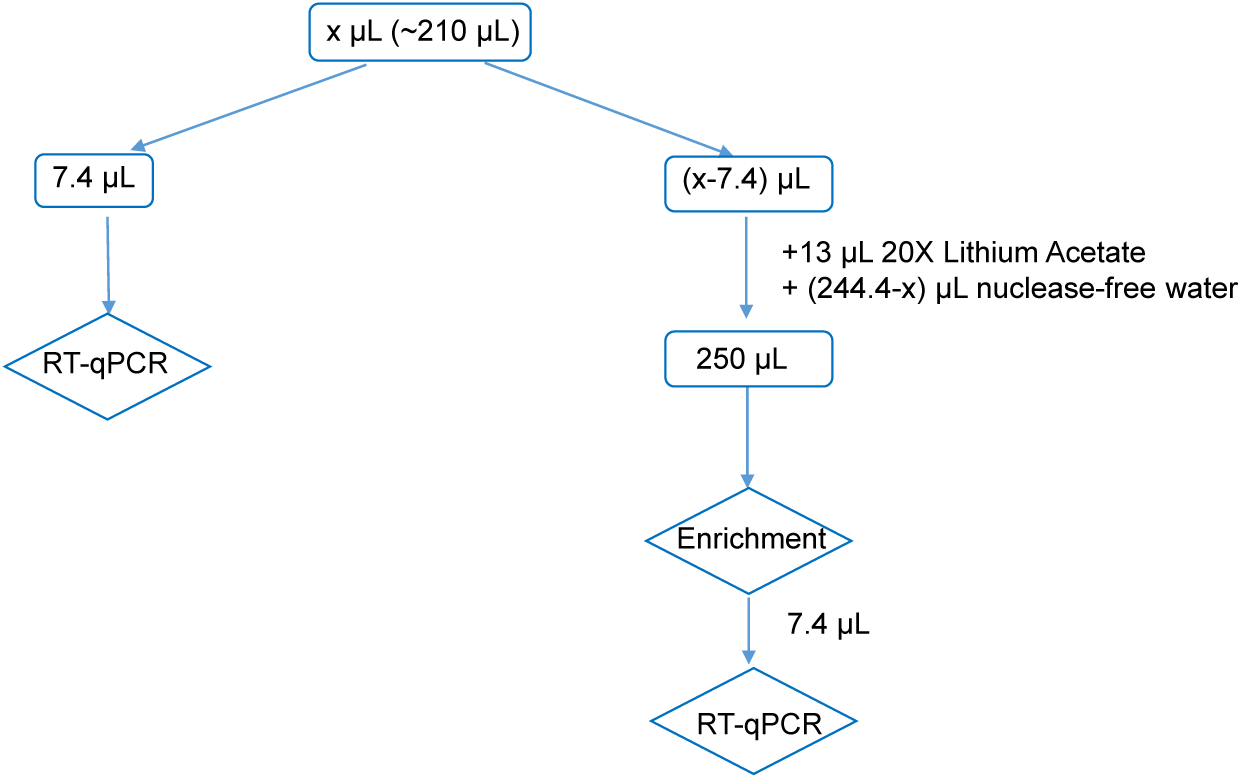
Flow chart of the integration experiments with the 430 genome copies (NTCs as well)

**Table S3:** Non-template controls in Integration tests. Left: these samples were collected from the 7.4 µL aliquots before enrichment (left side of Figure S3). Right: these samples were collected after enrichment (right side of Figure S3). All NTCs were negative.

| NTC # | Ct value | NTC # | Ct value | NTC # | Ct value | NTC # | Ct value |
| --- | --- | --- | --- | --- | --- | --- | --- |
| 1 | ND | 9 | ND | 1 | ND | 9 | ND |
| 2 | ND | 10 | ND | 2 | ND | 10 | ND |
| 3 | ND | 11 | ND | 3 | ND | 11 | ND |
| 4 | ND | 12 | ND | 4 | ND | 12 | ND |
| 5 | ND | 13 | ND | 5 | ND | 13 | ND |
| 6 | ND | 14 | ND | 6 | ND | 14 | ND |
| 7 | ND | 15 | ND | 7 | ND | 15 | ND |
| 8 | ND |  |  | 8 | ND |  |  |
| Total detection rate: |  | 0/15 (0%) |  | Total detection rate: |  | 0/15 (0%) |  |

### Enrichment

#### RT-qPCR

**Figure S4:**
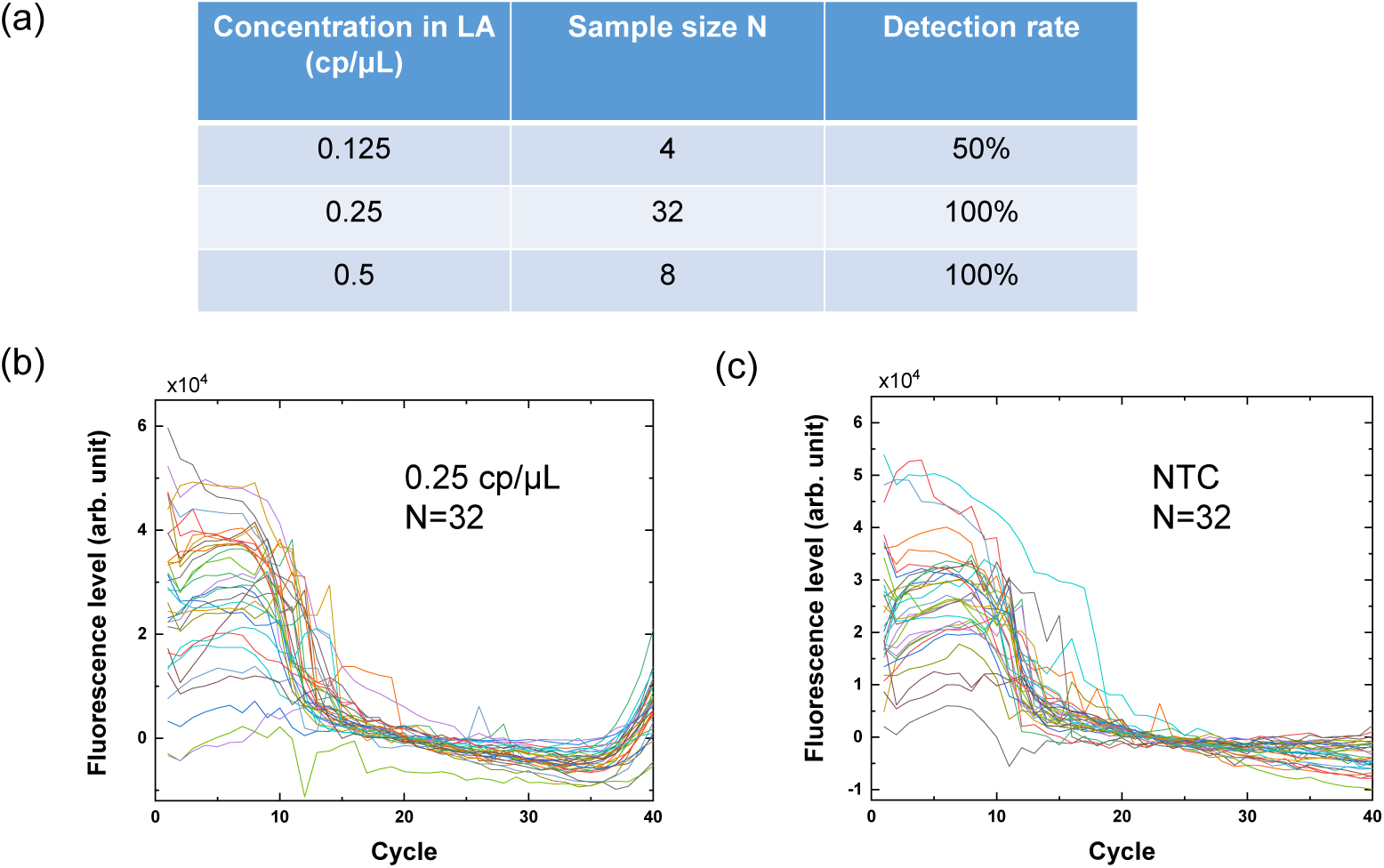
Fast RT-qPCR. (a) Positive detection rate of SARS-CoV-2 RNA in LA buffer at different concentrations. (b) Amplification curves (N=32) of 0.25 cp/ µL SARS-CoV-2 RNA in LA buffer. (c) Amplification curves (N=32) of non-template control in LA buffer.

**Figure S5:**
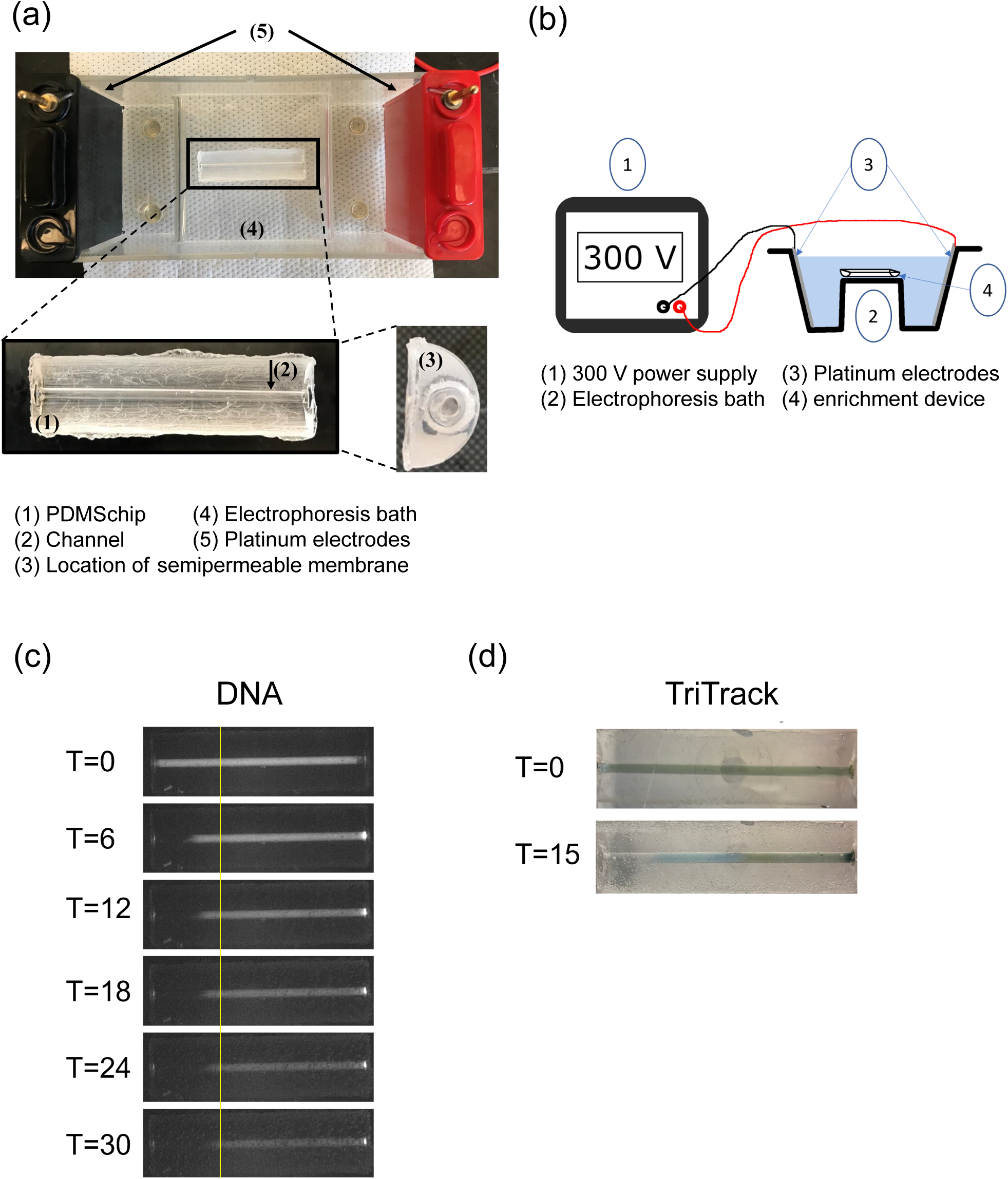
Nucleic acid enrichment by electrophoresis. (a) Photographs of the enrichment device and the electrophoresis bath used, in which the key elements are highlighted. (b) Schematic of the key elements of the enrichment process and their connections. (c) Enrichment of DNA 1 kb ladder stained with SYBR Safe dye. (d) Enrichment of TriTrack dye alone.

**Figure S6:**
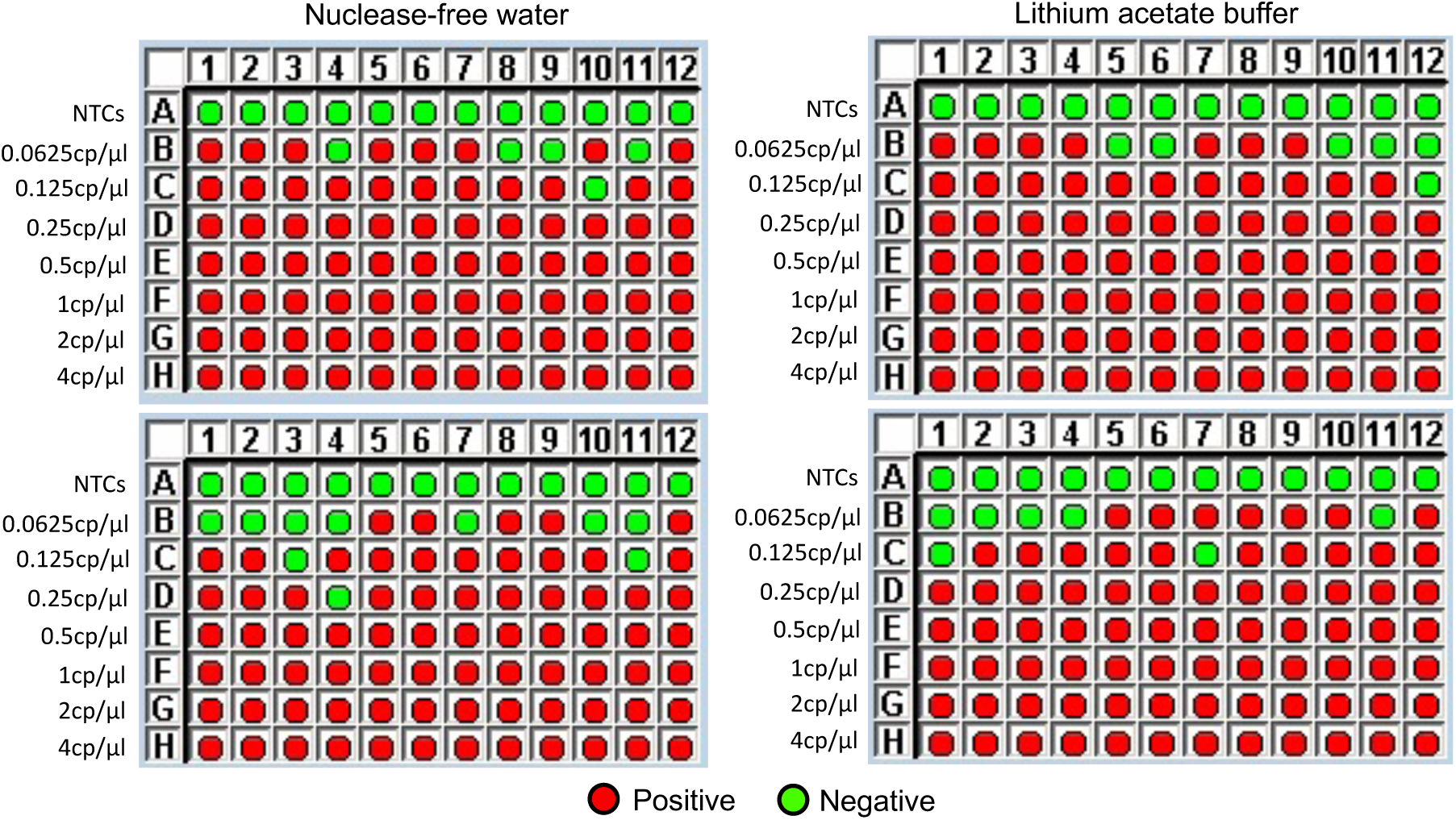
Results of RT-qPCRs utilized to generate the standard curves for nuclease-free water and lithium acetate.

**Figure S7:**
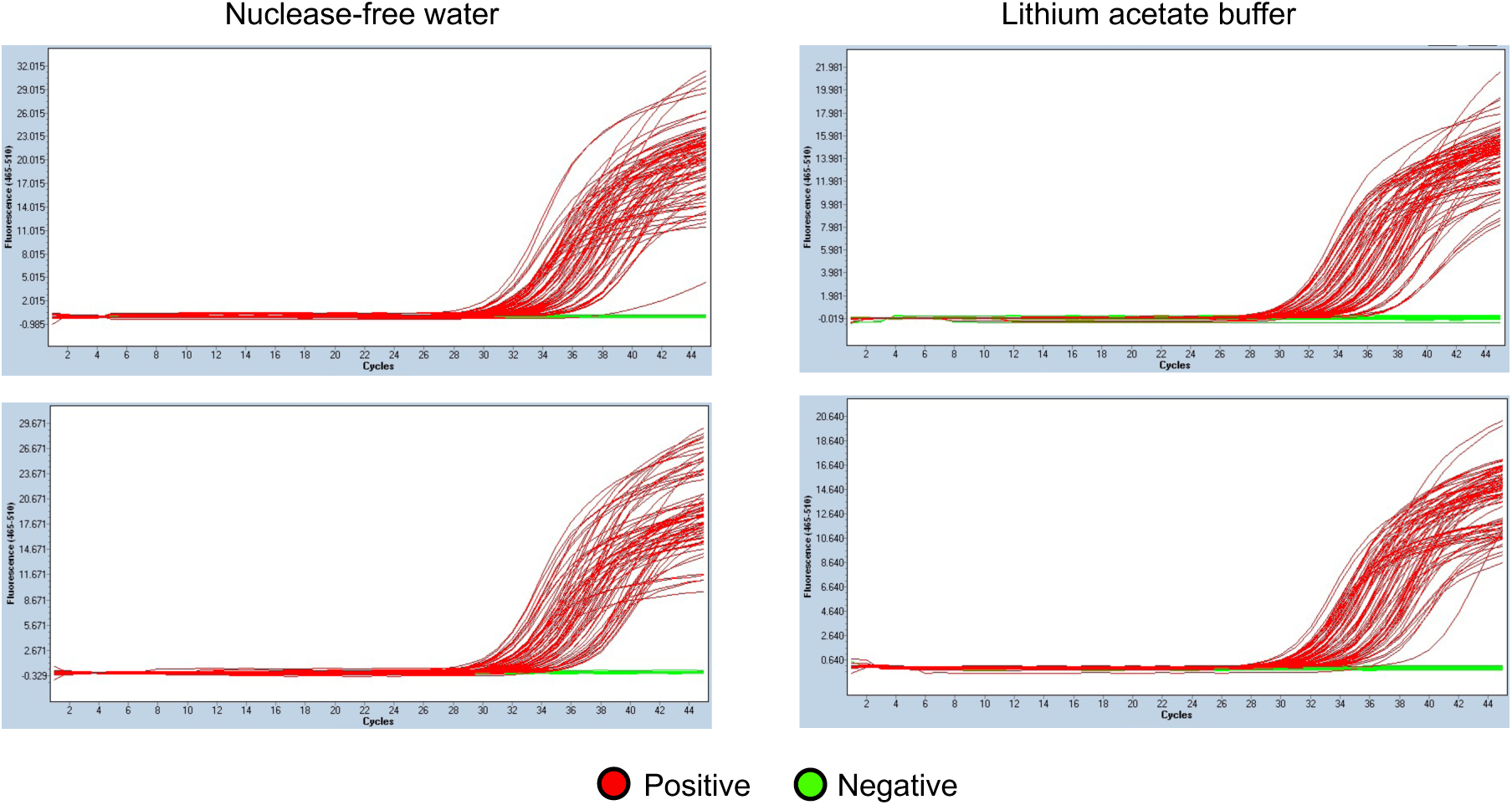
Amplification curves of RT-qPCRs utilized to generate the standard curves for nuclease-free water and lithium acetate.

